# Do People Reduce Attempts At Lifestyle Change After Initiating Weight Loss Medication? A Review Of Trial Evidence

**DOI:** 10.1101/2024.01.12.24301166

**Authors:** Megan Macfarlane, David Cooper, Alison Avenell

**Affiliations:** Health Services Research Unit University of Aberdeen Foresterhill Aberdeen UK AB25 2ZD

**Keywords:** Compensatory behaviours, anti-obesity medication, orlistat, liraglutide, semaglutide, low-fat reducing diet

## Abstract

Anti-obesity medications (AOMs) are used with lifestyle change support for weight management for adults with obesity. We investigated whether initiation of AOMs appeared to reduce attempted lifestyle changes. We searched for orlistat, liraglutide, semaglutide, and low-fat reducing diet (LFRD) trials in a trial register, Medline, Embase and systematic reviews from 1990 to June 2023. Weight changes in placebo groups of AOM trials with lifestyle interventions were compared to weight changes in LFRD trials. 16 orlistat, 7 liraglutide, 5 semaglutide, and 7 LFRD trials were included. Weight loss in orlistat placebo groups was 1.1kg less than LFRDs, a statistically non-significant difference, where last observation carried forward values were used for missing data in trials. Adjustment with baseline observation carried forward found 2.6kg less weight lost, a significant difference. Last observation carried forward data for orlistat trials providing completer data found a 0.2kg difference.

Liraglutide placebo groups had 2.1kg less weight lost than predicted from LFRD trials. Semaglutide placebo groups had 2.0kg less weight loss.

Compensatory reductions in changes to lifestyle behaviours may result following AOM initiation, but this needs to be explored further. Variability in trial design and study populations may contribute to the differences in weight change found.

## INTRODUCTION

Adults with obesity are supported to lose weight through changing diet and physical activity (PA) with behavioural interventions^1,2^. Where the target weight is not achieved or maintained, additional support can be provided through the initiation of anti-obesity medications (AOMs) ^1^. Currently, several AOMs have been approved for use by pharmaceutical regulatory agencies, including orlistat^3–5^, liraglutide^6–8^ and semaglutide^9–11^. Orlistat is a lipase inhibitor, blocking the function of gastrointestinal lipases and thus blocking the absorption of fat^3–5^. Liraglutide and semaglutide are both glucagon-like peptide-1 (GLP-1) agonists, also used by people with type 2 diabetes, which work by increasing insulin release and inhibiting glucagon release. However, GLP-1 agonists have also been found to delay gastric emptying and reduce appetite via central control, hence, assisting with weight loss^7,8,10–12^.

AOMs should be prescribed alongside a reduced calorie diet and modifications to PA, where possible, to maximise weight loss¹, meaning initiation of AOMs may promote healthy behaviours. It has, however, been suggested that compensatory behaviours may occur, with users tending to substitute a healthy lifestyle for taking a drug^13,14^. There are currently no studies assessing evidence of compensatory behaviours following AOM initiation. However, a recent scoping review^15^ looked for a relationship between initiation of lipid lowering drugs (LLD) and/or anti-hypertensive drugs and healthy diet in observational literature. Like AOMs, it is recommended that users of LLDs and anti-hypertensive drugs use them in addition to lifestyle changes to lower their cardiovascular disease risk factors further^16,17^. The review found that in 9 of the 17 included studies, use of LLDs or anti-hypertensive drugs was not always associated with improvements in diet^15^, suggesting lifestyle modification advice was either not received or was not acted upon by individuals starting these drugs. Similarly, a cohort study carried out between 2000 and 2013 on lifestyle behaviours following the initiation of anti-hypertensives and statins^18^ found initiators of the medications had a larger increase in BMI (difference of 0.19kg/m²) and were more likely to reduce PA (difference of −0.09 hours/day) compared with non-initiators. However, initiation did result in a reduction in alcohol consumption and higher likelihood of stopping smoking. It is possible that this increase in BMI resulted from replacing a healthier lifestyle with medication use. However, smoking cessation is associated with subsequent weight gain that cannot be reduced by PA alone^19^. Regardless, initiators may still be less healthy than their non-initiator counterparts. Statin use is associated with muscle complaints^20^ which might make it more difficult for some initiators to take part in PA^18^.

In 2005, Esposito and Giugliano^13^ commented on placebo participants limited weight loss during a randomised controlled trial (RCT) for a now unavailable AOM, rimonabant, implying they did not follow the prescribed dietary deficit of 600 kcal/d. In response, Brown and Avenell^14^ added that similar limited weight loss can be seen in placebo groups of orlistat RCTs. It was suggested that the participants of drug RCTs do not follow the dietary advice provided to participants to the same degree as those in dietary intervention RCTs, however, it was recognised that the higher BMI in participants of AOM RCTs may mean they may have an element of treatment resistance. More recently, Willet also described issues regarding behavioural compensation following initiation of AOMs^21^. A comparison of weight loss seen in AOM trials compared with non-drug diet trials is therefore warranted to see if this effect is present.

## METHODS

The search strategy, study eligibility criteria and statistical approaches of this systematic review were predefined in an unpublished research protocol.

### Study selection

Included trials were RCTs with a weight loss intervention for adults (age ≥18 years) with obesity (mean body mass index (BMI) at baseline ≥30kg/m²) with follow-up of at least 12 months (68 weeks for semaglutide) from randomisation. Interventions included were (1) orlistat dose of 120mg three times daily, (2) liraglutide dose 3.0mg subcutaneously once daily, (3) semaglutide dose 2.4mg subcutaneously once weekly or (4) low-fat reducing diet (LFRD) advice. Drug RCTs were compared to placebo. Included drug trials included advice on hypocaloric diets to all trial groups, but did not include those with meal replacements. For orlistat RCTs, advice regarding a LFRD (stating low-fat and/or less than 30% of dietary intake from fat), as per orlistat prescribing guidance^3^, must have been provided to all participants. LFRD intervention RCTs were compared with no or minimal intervention and did not describe providing physical activity (PA) advice, allowing for comparison of weight change to orlistat RCTs where PA advice was often not described.

### Literature search

The reference list of a systematic review on the efficacy of anti-obesity medications ^22^ (last date of search strategy March 23, 2021) was searched for relevant orlistat, liraglutide and semaglutide RCTs. Additionally, a database search of Medline (Ovid) and Embase (Ovid) for drug weight loss RCTs from 1990 to present (January 06, 2023 for orlistat; June 19, 2023 for liraglutide; June 20, 2023 for semaglutide) was carried out.

A rapid search of the reference lists of previous systematic reviews^23–25^ was conducted for LFRD RCTs. Additionally, a database search of the University of Aberdeen Health Services Research Unit’s obesity trials’ register for low-fat weight loss trials was undertaken on January 13, 2023. This database, of all adult weight loss trials with a follow-up of at least one year, contains relevant searches of Medline, Embase and the Cochrane Register of RCTs and had been continuously updated from 1966 until 2017. Finally, a brief database search for LFRD RCTs from 2017 to January 2023 using search strategy for RCT and obesity and low fat.tw or low-fat.tw was completed. Duplicates were removed, and titles and abstracts were screened and excluded if not relevant. Full texts were independently analysed by two authors (AA and MM) to produce the final list of included weight loss trials.

### Data extraction

Data were extracted regarding the (1) inclusion and exclusion criteria of each trial, (2) location of the study, (3) source of funding, (4) run-in period, (5) the number of contacts during the first year of the study, (6) advice given and (7) by whom, and (8) the methods of statistical analysis. The baseline characteristics of participants, including (a) mean age, (b) sex, (c) mean BMI, (d) co-morbidities, (e) ethnicity, and (f) socioeconomic status were also extracted. Data were extracted by MM and verified by AA with differences resolved through discussion.

For orlistat and LFRD RCTs, the mean weight change in kilograms (kg) and standard deviation (SD) at 12 months were extracted. Last observation carried forward (LOCF) data were used when provided if no other data for all participants randomised were available. Where trials also provided separate data for completers only, this was extracted and analysed separately.

Most liraglutide RCTs presented weight change as a percentage at 12 months and, as such, data were extracted and analysed in this form. Semaglutide RCTs also presented weight change as percentage change at 68 weeks and these data were extracted for analysis. Few liraglutide and semaglutide trials presented measures of variability and it was decided these values would not be estimated, thus meta-analysis was not possible for these trials. The weighted mean for percentage weight change for these trials was therefore calculated and used to estimate weight change in kg. Where data were presented in a different format to those required, relevant formulae were used as previously described elsewhere^24^.

Data for the number of participants randomised and the number of completers (on-drug) were extracted to calculate the number of dropouts. Where the number of completers at 12 months (68 weeks for semaglutide) was not stated, the number of completers at the end of the study was used. Where there were several intervention arms, dropouts for relevant arms were combined by addition of data.

### Data analysis and reporting

Meta-analysis was used to analyse pooled data, involving comparison groups from included studies. Orlistat intervention groups were compared to placebo groups and LFRD intervention groups were compared to control. Review manager version 5.4^26^ was used to conduct meta-analyses to compare interventions. For weight, a random-effects model was used since high heterogeneity between trials was expected. Data were analysed at 12 months to allow a common follow-up time. This was also the most common follow-up time, allowing for maximal data for comparison. A continuous inverse variance method was used to calculate the mean difference for weight change data and a dichotomous inverse variance method was used to calculate the mean difference for dropout data. A dichotomous Mantel-Haenszel method was used to calculate the risk ratio (RR). Heterogeneity was tested using the I² statistic with an I² value greater than 50% suggesting moderate heterogeneity^27^.

To assess the effect AOM initiation has on lifestyle behaviours, the pooled mean of the weight changes in the placebo groups in orlistat, liraglutide, and semaglutide RCTS were compared to the mean difference in weight change between the LFRD groups and control.

Statistical methods for analysis of weight differences - the pooled mean, pooled variance, and confidence intervals (CI) for the orlistat and placebo groups were calculated to allow for comparison to the LFRD RCTs. The pooled mean and variance were calculated using the respective formulae 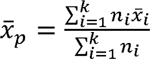, and 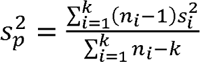, where:

*n*_i_ are the number of observations in trial *i* receiving the respective treatment

*x*^^^_i_ is the mean weight change in trial *i* for the respective treatment

*S^2^_i_* is the variance in trial *i* for the respective treatment

*k* are the number of trials of the respective treatment

The confidence interval for the two groups is obtained as xp ±1.96 √s^2^_p_/n_p_ where: the total number of observations, *n*_p_ = Σ^k^_i=1_ n_i_

The meta-analysis mean difference was used to obtain the estimated effect size for the LFRD with the standard error estimated by the difference in the upper and lower limits of the confidence interval divided by 3.92. These were then compared using a two sample independent t-test to the mean and standard deviation obtained from the pooled means and variances of the respective orlistat and placebo trials to make a formal statistical comparison.

Baseline observation carried forward adjustments – a post hoc adjustment of weight data from LOCF to baseline observation carried forward (BOCF) for participants that dropped out of the orlistat trials was completed for trials that reported separate data for completers. The method for this adjustment is described elsewhere^28^.

## RESULTS

### Search results

Figure S1 in the online supplement presents the study selection flow diagram.

### Study characteristics

The descriptions of drug studies included in this review are summarised in the Table S1 in the supplement. 12/16 orlistat RCTs, 1/7 liraglutide RCTs and 0/5 semaglutide RCTs described a run-in period prior to randomisation. 10/16 orlistat RCTs, 7/7 liraglutide RCTs and 5/5 semaglutide RCTs reported giving advice regarding PA to participants. No RCTs provided a PA programme for participants to attend. The number of contacts by 12 months (68 weeks for semaglutide) ranged from 11 to 20 in orlistat RCTs, 12 to 23 in liraglutide RCTs and 12 to 21 in semaglutide RCTs. 8/16 orlistat RCTs, 7/7 liraglutide RCTs, and 5/5 semaglutide RCTs reported the advice and counselling received by participants was from a dietician or similarly qualified healthcare professional.

The descriptions of each included LFRD RCT are summarised in the Table S2 in the supplement. LFRD RCTs did not describe run-in periods prior to randomisation or describe providing PA advice to participants. Participants in the control groups were told to not change their dietary or PA patterns, other than one RCT^29^ where the control group were told to try to lose weight, however, they were given no formal counselling. The number of contacts by 12 months of the studies ranged from 4 to 32. Where details regarding who provided counselling advice were reported, RCTs had registered dieticians providing counselling.

The baseline characteristics of the participants of each drug RCT included in this review are summarised in the Table S3 in the supplement. The age ranges of participants at baseline were similar between the drug RCTs. All drug RCTs tended to recruit more female than male participants. Participants in orlistat RCTs (means 33kg/m²-38kg/m²) tended to have a lower mean baseline BMI compared to participants in liraglutide RCTs (means 35kg/m²-40kg/m²) and semaglutide RCTs (means 36kg/m²-39kg/m² apart from one RCT^30^ where only participants of Asian ethnic origin were included, and the mean baseline BMI was 32kg/m²). 8/16 of orlistat RCTs, 4/7 liraglutide RCTs and 5/5 semaglutide RCTs recruited participants due to their co-morbidities. Where details regarding the ethnicity of participants were provided, the majority of participants in each RCT were white. Details regarding the socioeconomic status of participants were not provided in any of the drug RCTs.

The baseline characteristics of participants in LFRD RCTs are summarised in Table S4 in the supplement. The age ranges of participants were similar to those in the drug RCTs. 3/7 RCTs included more female than male participants, with one of those trials including only female participants. The mean baseline BMI ranged from 29 kg/m² to 35 kg/m². The BMI for the RCT^31^ with BMI below 30kg/m² was for completers of the intervention, not for those randomised. It was therefore presumed that the BMI of participants randomised would be ≥30kg/m² and therefore included in this review. 5/7 RCTs recruited participants due to their co-morbidities. Where details regarding the ethnicity of participants were provided, the majority of participants in each RCT were white. Only one RCT reported on the socioeconomic status of participants.

All drug RCTs were sponsored by the drug manufacturer: orlistat RCTs were sponsored by Roche, and both liraglutide and semaglutide RCTs were sponsored by Novo Nordisk. One LFRD RCT^29^ was funded by Astra-Merck. Another LFRD RCT^32^ was funded by the insurance company “Vital Friskvern”. The remaining LFRD RCTs were not funded by commercial companies.

### Weight change

#### Orlistat: all included RCTs

The mean weight reductions in those randomised to the orlistat intervention groups were always larger than those in the placebo groups. All placebo groups lost weight. Figure 1 shows the forest plot for meta-analysis of orlistat RCTs included 10,181 participants in which data for weight change at 12 months were available. The mean difference [95%CI] for weight change between the orlistat intervention groups and the placebo/control groups was −3.0kg [-3.5, −2.6]. Heterogeneity between the studies was 63% suggesting moderate heterogeneity between the trials.

The weighted mean weight change was calculated individually for the orlistat groups and the placebo groups. For the orlistat intervention group, the mean [95%CI] weight change was −7.0kg [-7.2, −6.8] (Figure 1, blue boxes). The mean [95%CI] weight change in the placebo group was −3.7kg [-3.8, −3.5] (Figure 1, red boxes).

**Figure 1.**
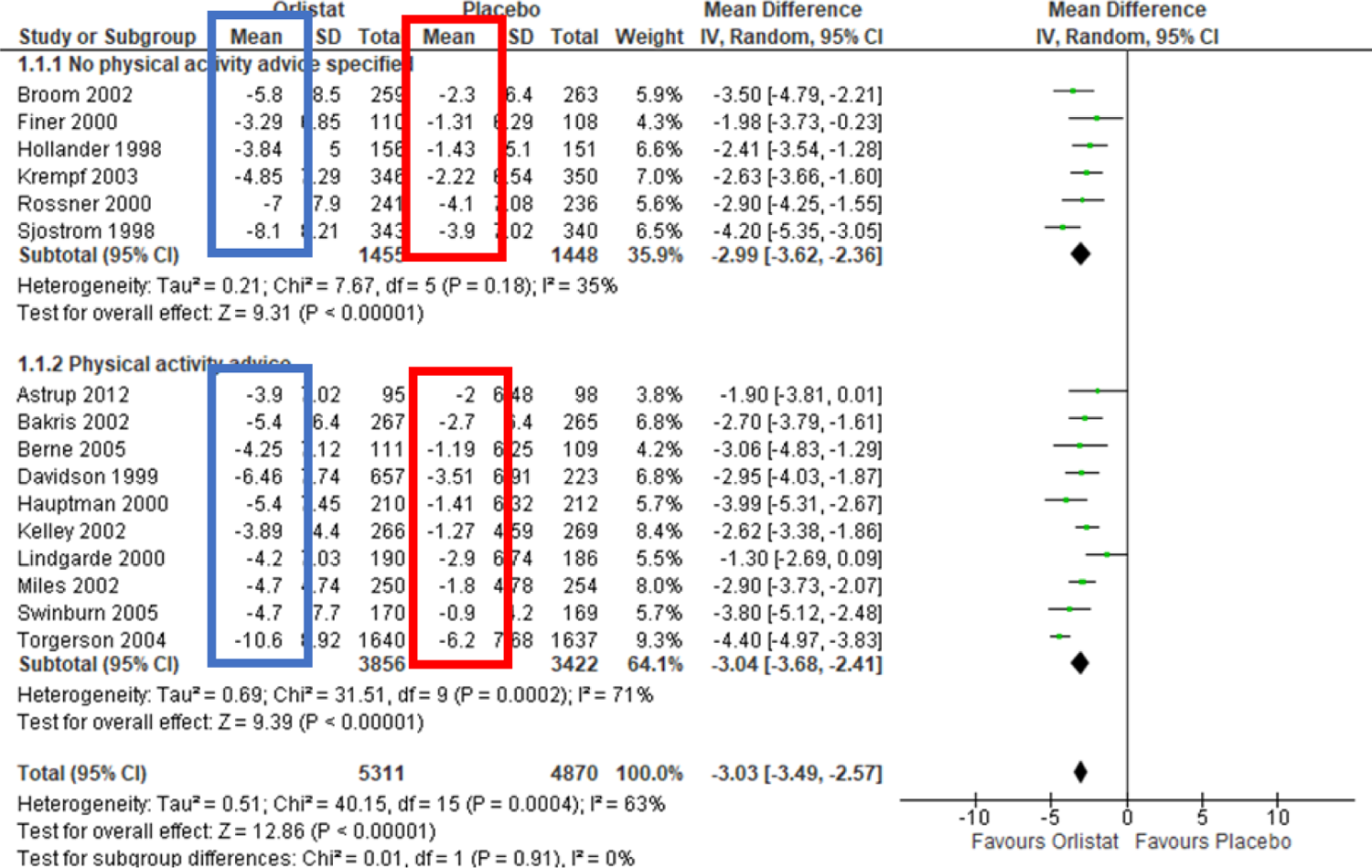
Mean difference weight change in kg for all orlistat trials versus placebo

#### LFRD

The mean weight reductions were always larger in the LFRD groups than the control. Figure 2 shows the forest plot for meta-analysis of LFRD RCTs included 1,008 participants in which data at 12 months were available. The mean difference [95%CI] for weight change between the LFRD groups and the control groups was −4.8kg [-6.2, −3.4]. Heterogeneity between the studies was 69% suggesting moderate heterogeneity between the trials.

**Figure 2.**
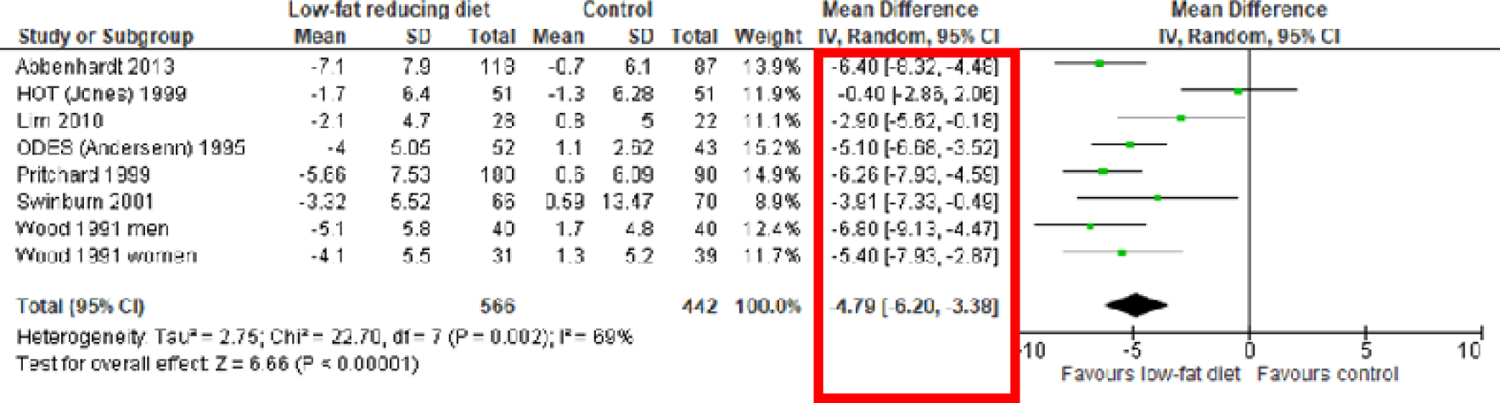
Mean difference weight change in kg for all LFRD trials versus control

#### Liraglutide

Table 1 shows the weight change (%) of those randomised in liraglutide RCTs. Those randomised to liraglutide always had a larger weight change than those randomised to placebo. The weighted mean weight changes for 3,511 participants randomised to liraglutide 3mg per day and 2,093 participants randomised to placebo in the liraglutide RCTs were calculated. Those randomised to liraglutide lost, on average, 7.8% of their initial body weight. Those randomised to placebo lost, on average, 2.5% of their initial body weight. When the weighted average baseline weight was calculated and used to calculate weight change in kg, those randomised to liraglutide lost an estimated 8.2kg compared to those randomised to placebo who lost an estimated 2.7kg.

**Table 1.**
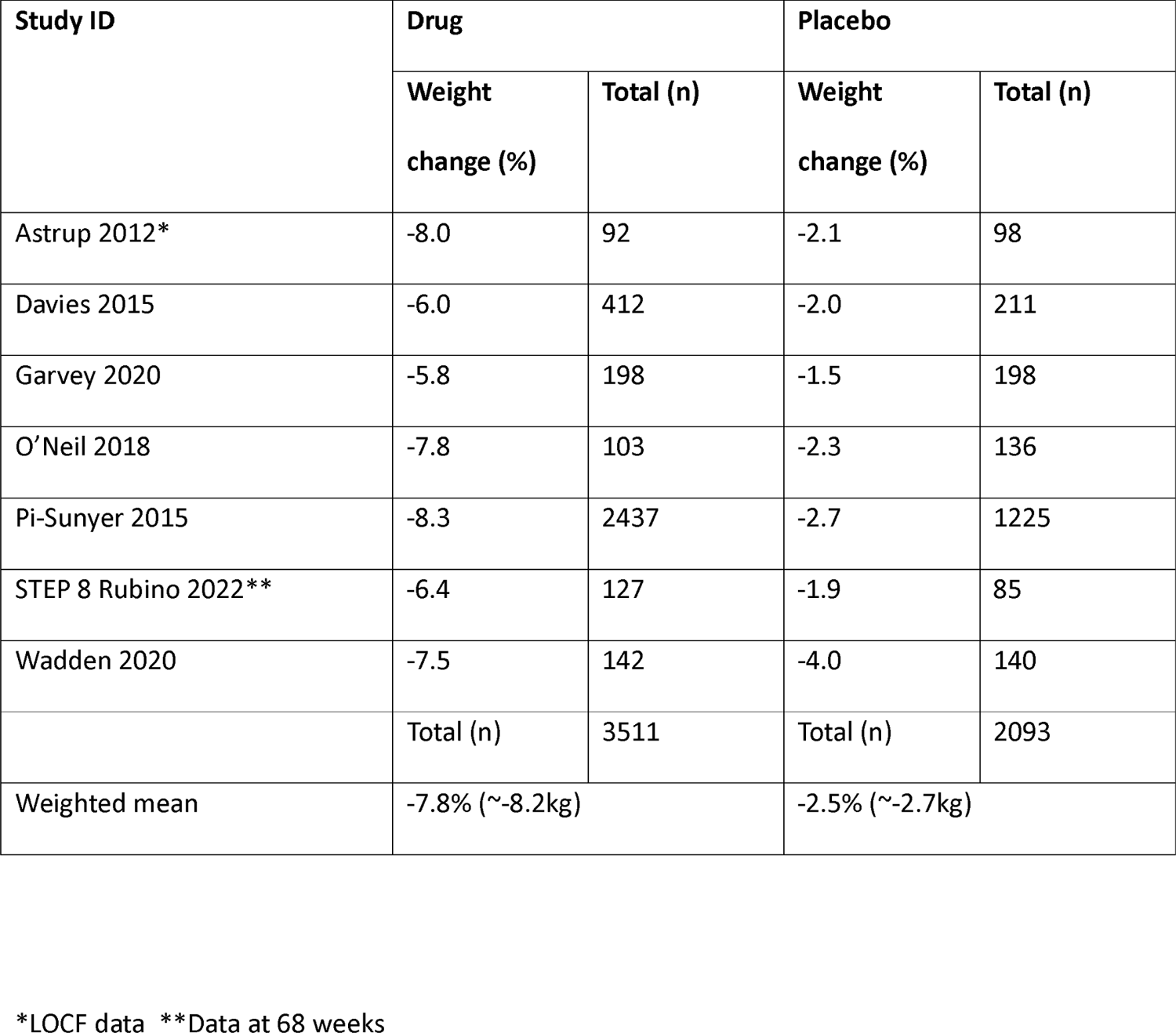
Weight changes in liraglutide trials.

#### Semaglutide

Table 2 shows the weight change (%) of those randomised in semaglutide RCTs. Those randomised to semaglutide always had a larger weight change than those randomised to placebo The weighted mean weight changes for 2,136 participants randomised to semaglutide 2.4mg per week and 1,333 participants randomised to placebo in the semaglutide RCTs were calculated. Those randomised to semaglutide lost, on average, 13.9% of their initial body weight. Those randomised to placebo lost, on average, 2.7% of their initial body weight. When the weighted average baseline weight was calculated and used to calculate weight change in kg, those randomised to semaglutide lost an estimated 14.3kg, compared to those randomised to placebo who lost an estimated 2.8kg.

**Table 2.**
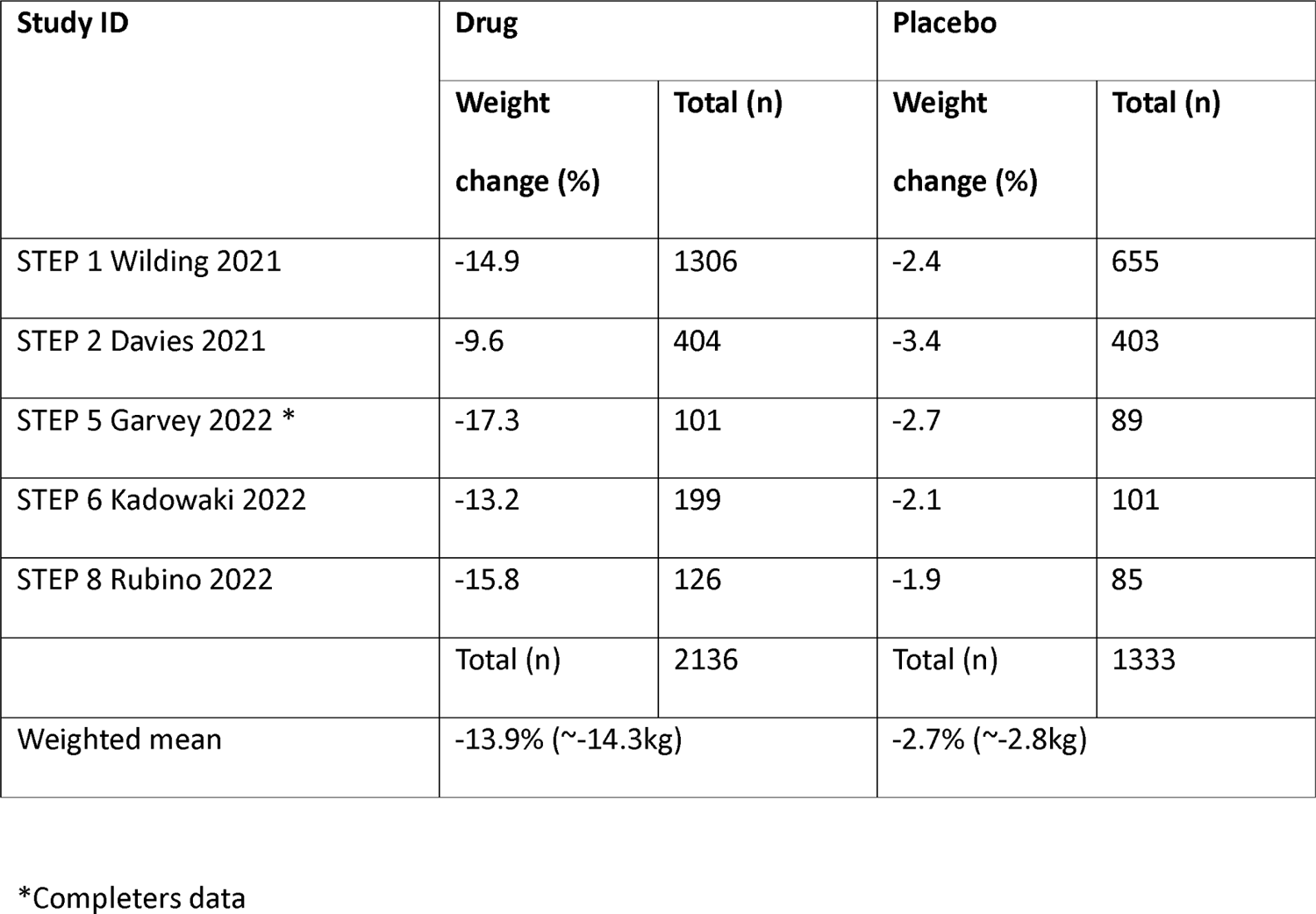
Weight changes in semaglutide trials.

### Attrition

#### Orlistat

Orlistat RCTs experienced significant numbers of dropouts throughout the trials. Meta-analysis of orlistat trials included 10,319 participants in which dropout data were available (Figure S2 in the supplement). The risk ratio [95%CI] for dropouts between the orlistat intervention groups and the

placebo groups was 0.8 [0.7, 0.9]. Participants randomised to the placebo group were more likely to drop out than the orlistat participants.

#### LFRD

Meta-analysis of LFRD RCTs included 911 participants in which dropout data were available (Figure S3 in the supplement). The risk ratio [95%CI] for dropouts between the LFRD groups and the control groups was 1.4 [1.1, 1.9]. Participants randomised to the LFRD group were more likely to drop out than the control participants.

#### Liraglutide

Meta-analysis of liraglutide RCTs included 5,686 participants in which dropout data were available (Figure S4 in the supplement). The risk ratio [95%CI] for dropouts between the liraglutide groups and the placebo groups was 0.8 [0.7, 1.0]. Those randomised to placebo were more likely to drop out than those randomised to liraglutide intervention.

#### Semaglutide

Meta-analysis of semaglutide RCTs included 3,583 participants in which dropout data were available (Figure S5 in the supplement). The risk ratio [95%CI] for dropouts between the semaglutide groups and the placebo groups was 0.8 [0.6, 1.0]. Those randomised to placebo were more likely to drop out than those randomised to semaglutide intervention.

### Adjustment to baseline observation carried forward (BOCF) for orlistat RCTs: trials presenting both completers and LOCF data

Orlistat trials that included data for completers only found a mean difference [95%CI] for weight change between the orlistat intervention groups and the placebo groups to be −3.3kg [-3.9, −2.6] (Figure 3). Following adjustments to BOCF for those who dropped out in these RCTs, the orlistat intervention groups had a mean [95%CI] weight change of −5.0kg [-5.4, −4.7] at 12 months. The mean [95%CI] weight change in the placebo group was −2.2kg [-2.5, −1.9].

**Figure 3.**
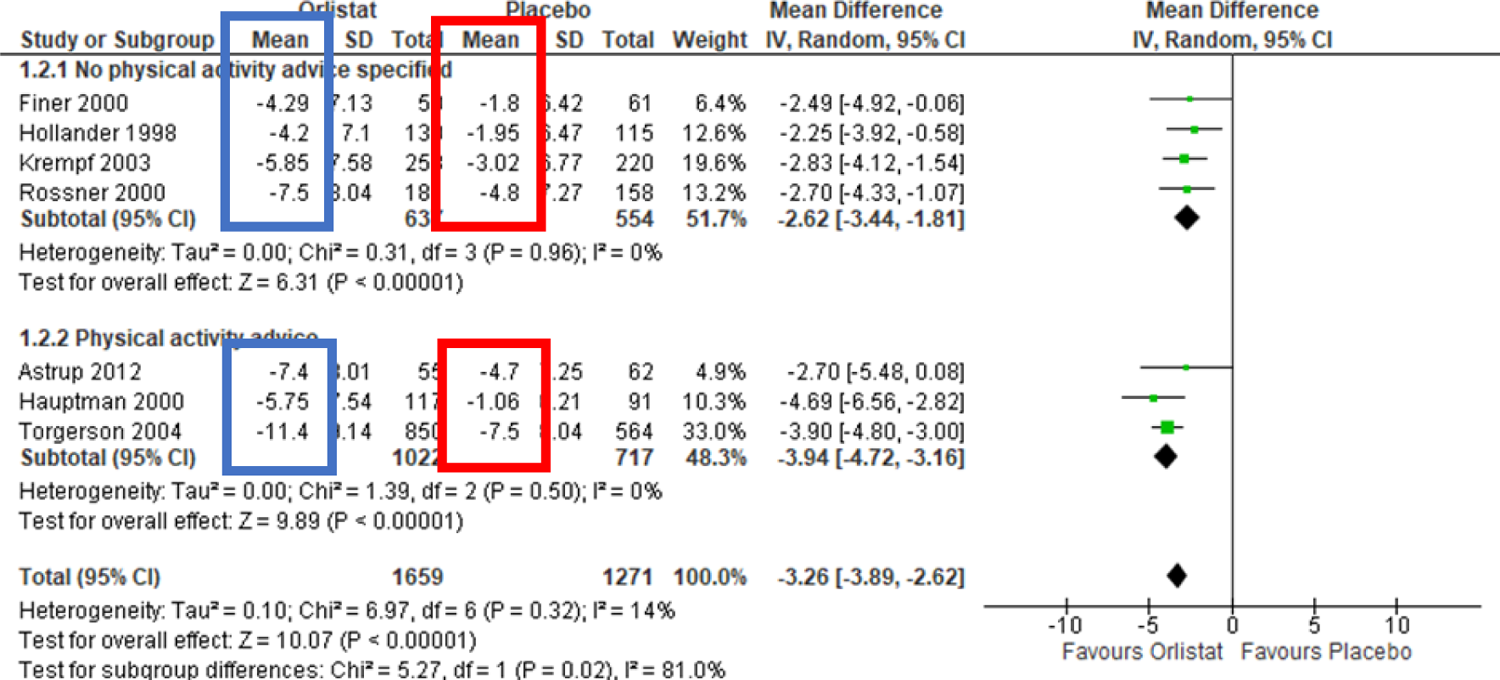
Mean difference weight change in kg for all orlistat trials versus placebo (completers only)

Where these trials only were analysed but using the original LOCF data, a mean difference [95%CI] for weight change between orlistat and placebo groups of −3.0kg [-3.9, −2.2] was found. The orlistat intervention groups had a mean [95%CI] weight change of −8.2kg [-8.5, −7.9]. The mean [95%CI] weight change in the placebo group was −4.6kg [-4.8, −4.3] (Figure S6 in the supplement).

### Analysis of weight difference between interventions

#### Orlistat vs LFRD

Comparison of the mean weight change in all the orlistat placebo groups (−3.7kg, 95%CI [-3.8, −3.5]) to the mean difference between the LFRD and the control groups (−4.8kg, 95%CI [-6.2, −3.4]) found a 1.1kg difference (95%CI [-0.3, 2.6] P=0.06).

Comparison of the mean weight change using the completer data for orlistat placebo groups (−2.2kg, 95%CI [-2.5, −1.9]) to the mean difference between the LFRD and the control groups (−4.8kg, 95%CI [−6.2, −3.4]) found a 2.6kg difference (95%CI [1.1, 4.0] p<0.001).

Comparison of the LOCF data for the orlistat placebo groups from trials which had provided completer data (−4.6kg, 95%CI [-4.8, −4.3]) to the mean difference between the LFRD and the control groups (−4.8kg, 95%CI [-6.2, −3.4]) found a 0.2kg difference (95%CI [-1.2, 1.7] p=0.38).

#### Liraglutide vs LFRD

Comparing the estimated mean weight change of the liraglutide placebo groups (−2.7kg) to the mean difference between the LFRD and the control groups (−4.8kg, 95%CI [-6.2, −3.4]) found a 2.1kg difference.

#### Semaglutide vs LFRD

Comparing the estimated mean weight change of the liraglutide placebo groups (−2.8kg) to the mean difference between the LFRD and the control groups (−4.8kg, 95%CI [-6.2, −3.4]) found a 2.0kg difference.

## DISCUSSION

Comparison of weight change seen in all orlistat placebo groups to the difference between the LFRD and control groups found a 1.1kg non-significant difference with less weight loss in the placebo groups. Adjusting values to BOCF for participants that dropped out in trials that also provided data for completers resulted in a 2.6kg significant difference between the placebo and the difference between the LFRD and control groups, with less weight loss seen in the placebo groups. Using LOCF data from trials that also provided completer data found a 0.2kg difference between weight change in the placebo groups and the difference between the LFRD and control groups, with less weight loss in the placebo groups.

In liraglutide RCTs, comparison of weight change seen in the placebo groups to the difference between the LFRD and control groups found a 2.1kg difference. Placebo groups lost less weight than the diet groups.

For semaglutide RCTs, comparison of weight change seen in the placebo groups to the difference between the LFRD and control groups found a 2.0kg difference with less weight loss in placebo groups.

As far as we are aware, this is the first review to assess compensatory lifestyle changes following the induction of AOMs. Although differences in weight loss seen between AOM RCTs and LFRD RCTs may be due to compensation, dissimilarities in the trial design and populations studied may also have explained differences.

12/16 orlistat RCTs included run-in periods prior to randomisation in their design. Run-in periods identify non-compliers and rule them out of randomisation. This is thought to decrease the number of dropouts during the study, improve compliance and hence, reduce the bias from incomplete data while improving the apparent effectiveness of the drug^33^. They may, however, cause problems regarding weight loss results. Most weight loss occurs early, and a rapid weight loss during the run-in period may result in less weight loss seen during the active intervention^34^. Although this may affect weight change results in orlistat RCTs, run-in periods were not included in the design of 6/7 liraglutide RCTs and 5/5 semaglutide RCTs, meaning the limited weight loss seen in placebo groups of these trials cannot be due to early weight loss prior to randomisation.

PA advice was reported as being given to participants in 10/16 orlistat RCTs, 7/7 liraglutide RCTs and 5/5 semaglutide RCTs, however, LFRD RCTs providing PA advice were excluded to allow for comparability to orlistat RCTs often with no PA advice described. Diet with PA produces more weight loss than diet alone^35^; meaning participants in most of the AOM RCTs may be likely to experience more weight loss than those in the LFRD RCTs. However, reported advice given to participants of RCTs was often vague and undetailed, the advice given may not have significantly affected the amount of weight loss experienced.

Baseline BMI was higher in orlistat RCTs than LFRD RCTs, and even more so in liraglutide and semaglutide RCTs. Varying evidence has been found regarding baseline BMI and the degree of weight loss experienced^36^. Features such as higher waist circumference, total body weight, fat-free mass, and muscle mass have been correlated to weight loss success^37^, meaning those in AOM RCTs may be more likely to experience weight loss success.

AOM RCTs included a higher ratio of female participants than LFRD RCTs. A systematic review and meta-analysis of RCTs found no significant gender differences^38^. However, due to RCTs providing limited details regarding interventions, it was not clear whether dietary prescription took account of body size in men and women. Regardless, gender differences between included RCTs should not significantly affect results.

Previous weight loss attempts of participants were not reported in any RCT. Participants with previous attempts at weight loss are typically less successful subsequently, with speculation that this is due to treatment resistance^39^. Since lifestyle attempts at weight loss are typically tried prior to AOM initiation, participants in AOM RCTs may have an element of treatment resistance and thus see less weight loss than expected.

AOM RCTs were all sponsored by their drug manufacturing company, increasing the risk of bias for these trials. Trials funded by pharmaceutical companies are more likely to report results favouring the drug^40^. As previously mentioned, reported information regarding lifestyle advice given to participants was very limited. It could be speculated that drug companies limit advice given to participants, which might enhance the drug effect. This would also result in less weight loss than expected, particularly for those in the placebo groups.

There were significant numbers of dropouts in AOM RCTs. Both liraglutide and semaglutide RCTs encouraged participants to continue to attend visits regardless of whether they were still taking the drug, reducing the amount of missing data. On the other hand, orlistat RCTs did not report encouraging participants to return, resulting in missing data that were subsequently substituted with LOCF data by the drug company. This method, however, assumes weight remains the same following withdrawal^28^. Participants typically have a period of weight regain following the initial weight loss^41^. Where participants drop out before weight is regained, LOCF data provided participants with a better result than if they were to remain in the trial. Adjustment to BOCF data was conducted for these participants^28^. BOCF would mean that all participants dropping out would return to their baseline weight, which also may not be the case. Both scenarios are not ideal^41^.

The difference in weight change between the LFRD group and control of LFRD RCTs was compared to the pooled mean weight change of the placebo groups of AOM RCTs. Conversely, a pooled mean for weight change for the LFRD intervention groups alone could have been used as comparison. This was calculated to be −4.9kg (95%CI [-5.4, −4.3]), 0.1kg more than data used, yielding a slightly greater difference in weight change seen than between placebo groups and LFRD groups. Another systematic review comparing low-fat diets to other diets, including usual diet ^42^, found a weighted mean difference between low-fat diet and usual diet of −5.43kg (95%CI [-7.29, −3.54]). Using these data would result in greater differences in weight change seen between placebo groups and LFRD groups. Our data, therefore, allowed for a more conservative approach in analysis.

The follow-up period was limited to 12 months, or 68 weeks for semaglutide RCTs, to allow for a common follow-up time. This timeframe reflects long-term weight loss, with weight loss interventions commonly seeing rapid weight loss early followed by a period of weight regain^34^. LFRD RCTs were required to include no PA advice to allow comparison to orlistat RCTs that often did not include PA advice. Several orlistat RCTs did, however, include PA advice and removing this from the exclusion criteria for LFRD RCTs would allow for more studies to be included in this group of trials. We only examined AOMs which have a licence in the UK. These analyses could be extended to other medications. We also did not examine drug trials where meal replacements were used, which would then be compared to trials of weight loss including meal replacements.

Further research is necessary to assess compensatory behaviours following the initiation of AOMs. One way of assessing this might be to conduct a 3-armed crossover RCT of drug and LFRD versus placebo and LFRD versus LFRD, where all participants could eventually receive the study drug. This would allow for comparison between groups while ensuring advice received by participants was the same. Cohort studies assessing lifestyle changes in relation to AOM initiation would also be helpful. Assessing the impact findings have on the economic evaluation of AOMs, where differences between drug and placebo groups are used, is essential. Finally, it would be important to conduct interviews with patients and doctors about their experiences with AOMs and their beliefs on how initiation affects lifestyle behaviours.

## CONCLUSION

Compensation for lifestyle measures may result following the initiation of AOMs. This could mean pharmacological management of obesity may not result in as much weight loss as expected with initiators gradually reverting to or continuing more of their unhealthy behaviours than expected. However, it may also be possible that participants of AOM RCTs have “treatment resistance” and hence, find it more difficult to lose weight through lifestyle behaviours. Regardless, for those starting anti-obesity medications, it is important to stress the importance of continuing caloric restriction and PA alongside the drug. Further studies are needed to assess these behaviours following the initiation of AOMs.

## ABBREVIATIONS

PA: physical activity

AOM: anti-obesity medications

GLP-1: glucagon-like peptide-1

LLD: lipid lowering drugs

RCT: randomised controlled trial

kcal: kilocalorie

BMI: body mass index

LFRD: low-fat reducing diet

kg: kilogram

SD: standard deviation

LOCF: last observation carried forward

RR: risk ratio

CI: confidence interval

BOCF: baseline observation carried forward

## Data Availability

All data used are available from the following papers: Abbenhardt et al. 2013 https://doi.org/10.1111/joim.12062, Anderssen et al. 1995. Oslo Diet and Exercise Study: a one year randomized intervention trial. Effect on hemostatic variables and other coronary risk factors. Nutrition Metabolism and Cardiovascular Diseases vol 5. Astrup et al. 2012 https://doi.org/10.1038/ijo.2011.158, Bakris et al. 2002 https://doi.org/10.1097/00004872-200211000-00026, Berne 2005 https://doi.org/10.1111/j.1464-5491.2004.01474.x, Broom et al. 2002 http://www.ncbi.nlm.nih.gov/pubmed/12296610, Davidson et al. 1999 https://doi.org/10.1001/jama.281.3.235, Davies et al. 2021 https://doi.org/10.1016/S0140-6736(21)00213-0, Davies et al. 2015 https://doi.org/10.1001/jama.2015.9676 Finer et al. 2000 https://doi.org/10.1038/sj.ijo.0801128 Garvey et al. 2022 https://doi.org/10.1038/s41591-022-02026-4 Garvey et al. 2020 https://doi.org/10.2337/dc19-1745 Hauptman et al. 2000 https://doi.org/10.1001/archfami.9.2.160 Hollander et al. 1998 https://doi.org/10.2337/diacare.21.8.1288 Jones et al. 1999 https://doi.org/10.1016/S0895-7061(99)00123-5 Kadowaki et al. 2022 https://doi.org/10.1016/S2213-8587(22)00008-0 Kelley et al. 2002 https://doi.org/10.2337/diacare.25.6.1033 Krempf et al. 2003 https://doi.org/10.1038/sj.ijo.0802281 Lim et al. 2010 https://doi.org/10.1016/j.numecd.2009.05.003 Lindgarde 2000 https://doi.org/10.1046/j.1365-2796.2000.00720.x Miles et al. 2002 https://doi.org/10.2337/diacare.25.7.1123 O Neil et al. 2018 https://doi.org/10.1016/S0140-6736(18)31773-2 Pi-Sunyer et al. 2015 https://doi.org/10.1056/nejmoa1411892 Pritchard et al. 1999. https://doi.org/10.1136/jech.53.5.311 Rossner et al. 2000 https://doi.org/10.1038/oby.2000.8 Rubino et al. 2022 https://doi.org/10.1001/jama.2021.23619 Sjostrom et al. 1998 https://doi.org/10.1016/S0140-6736(97)11509-4 Swinburn et al. 2005 https://doi.org/10.1111/j.1463-1326.2004.00467.x Swinburn et al. 2001 https://doi.org/10.2337/diacare.24.4.619 Torgerson et al. 2004 https://doi.org/10.2337/diacare.27.1.155 Wadden et al. 2020 https://doi.org/10.1002/oby.22726 Wilding et al. 2021 https://doi.org/10.1056/nejmoa2032183 Wood et al. 1991 https://doi.org/10.1056/NEJM199108153250703 All data produced in the present work are contained in the manuscript

## Funding

The Health Services Research Unit is funded by the Chief Scientist Office of the Scottish Health and Social Care Directorates.

### Acknowledgements

We thank Paul Manson, Information Specialist at the Health Services Research Unit, University of Aberdeen, for assistance with search strategies.

## ONLINE SUPPLEMENT

**Figure S1.**
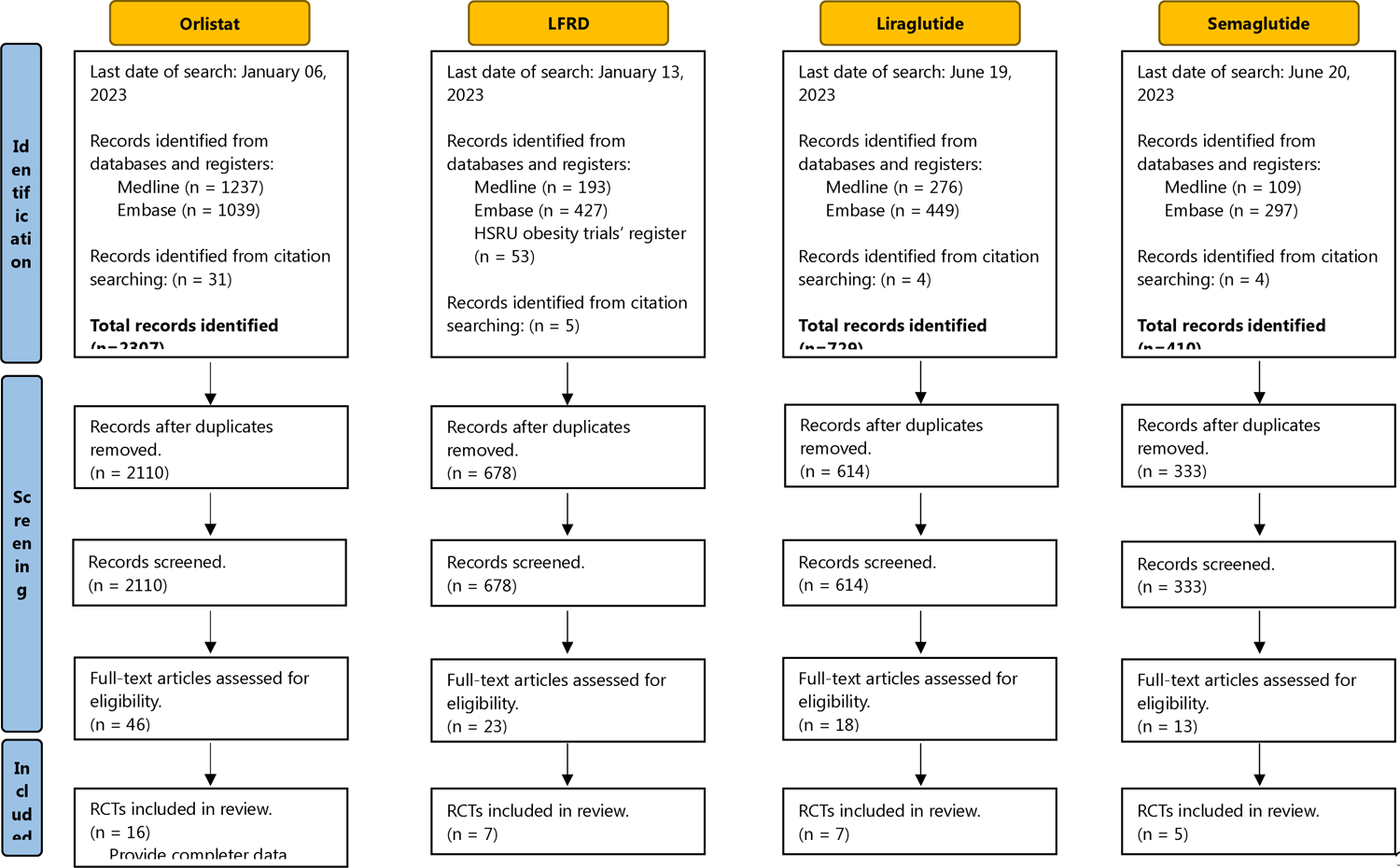
Modified PRISMA diagram for all included randomised controlled trials

**Figure S2.**
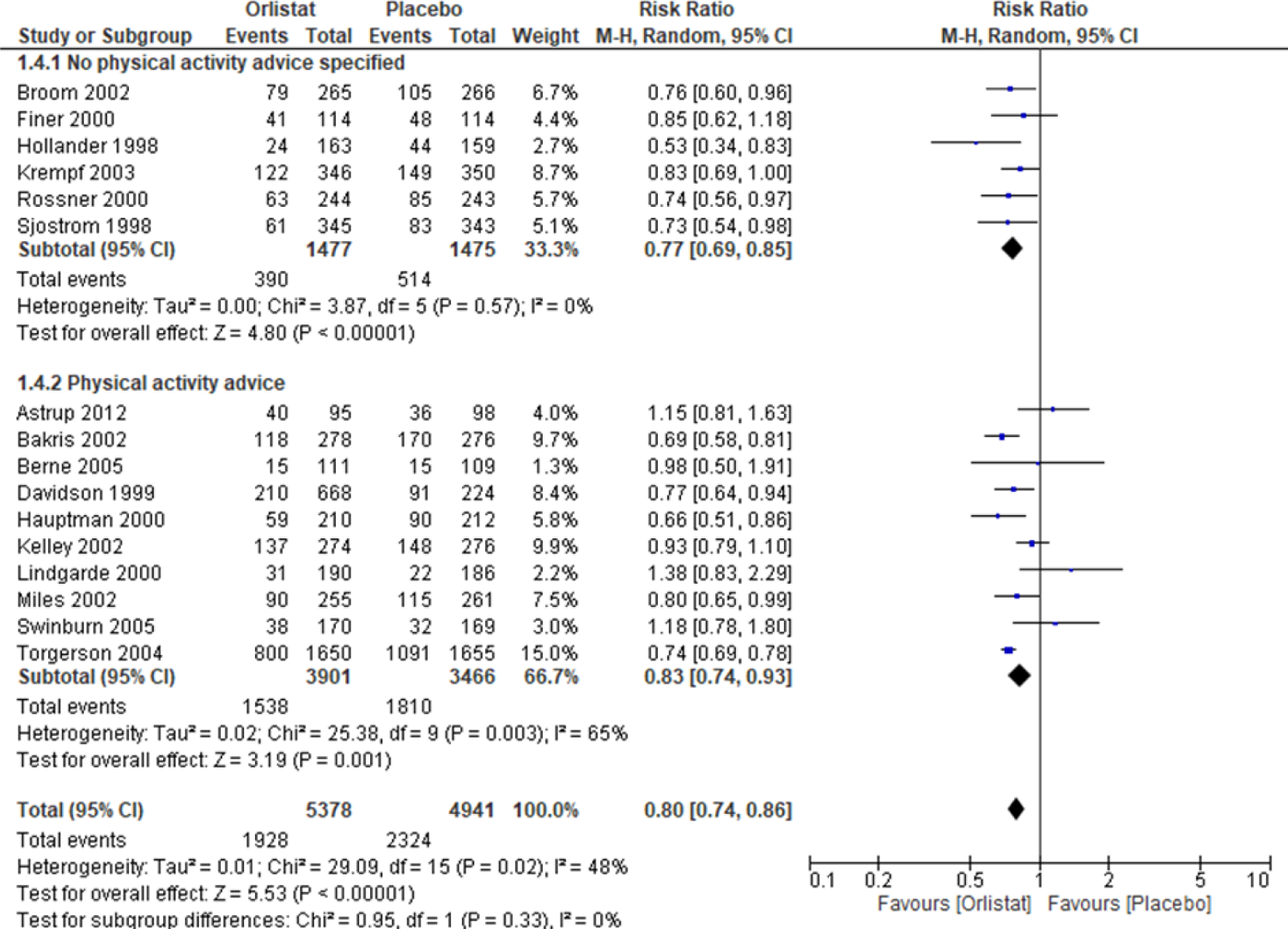
Forest plot for meta-analysis of the effect of orlistat versus placebo on the number of dropouts at 12 months follow-up

**Figure S3.**
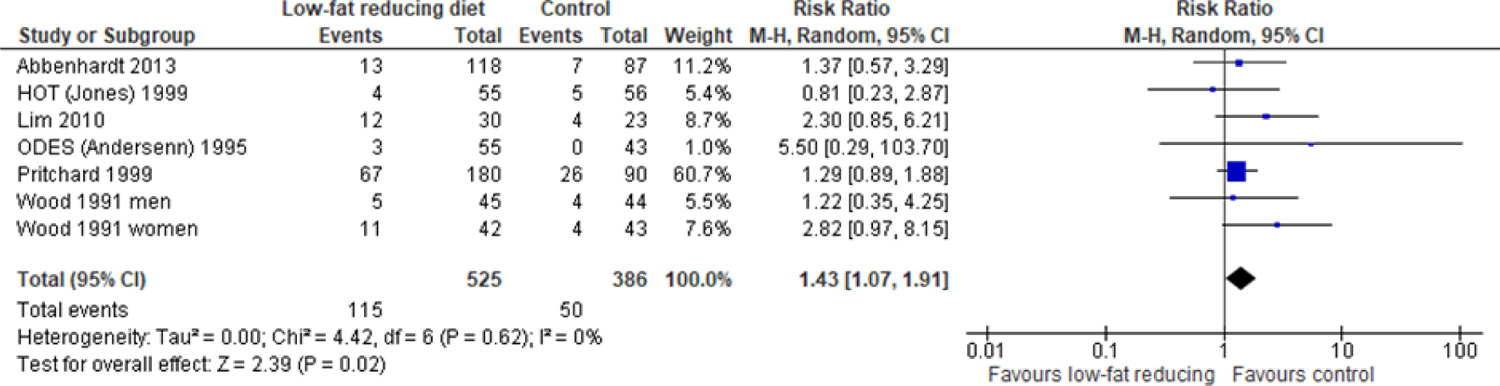
Forest plot for meta-analysis of the effect of LFRD versus control on the number of dropouts at 12 months follow-up

**Figure S4.**
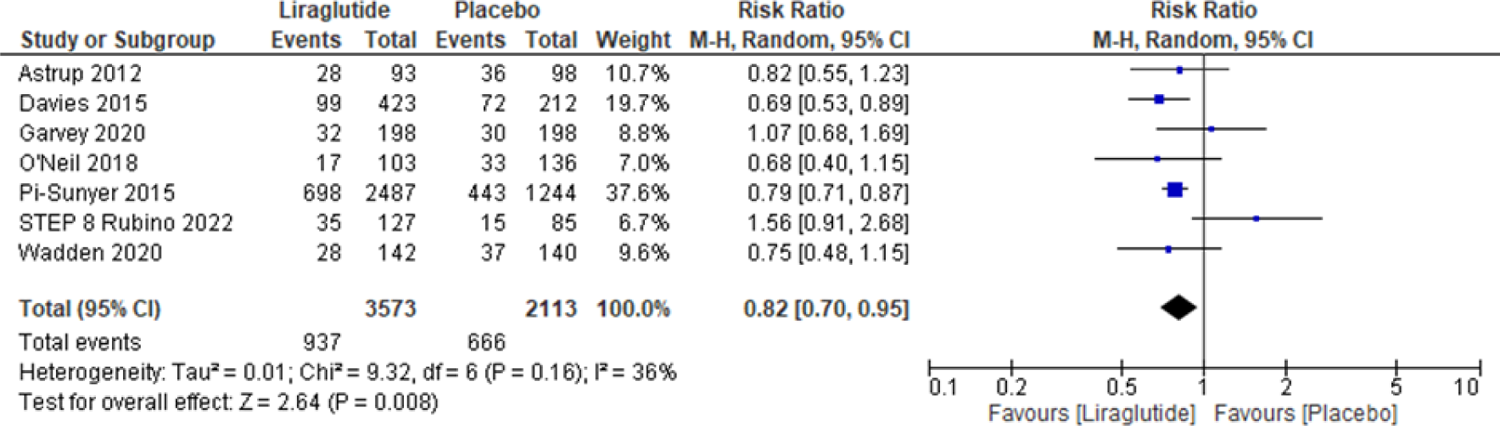
Forest plot for meta-analysis of the effect of liraglutide versus placebo on the number of dropouts at 12 months follow-up

**Figure S5.**
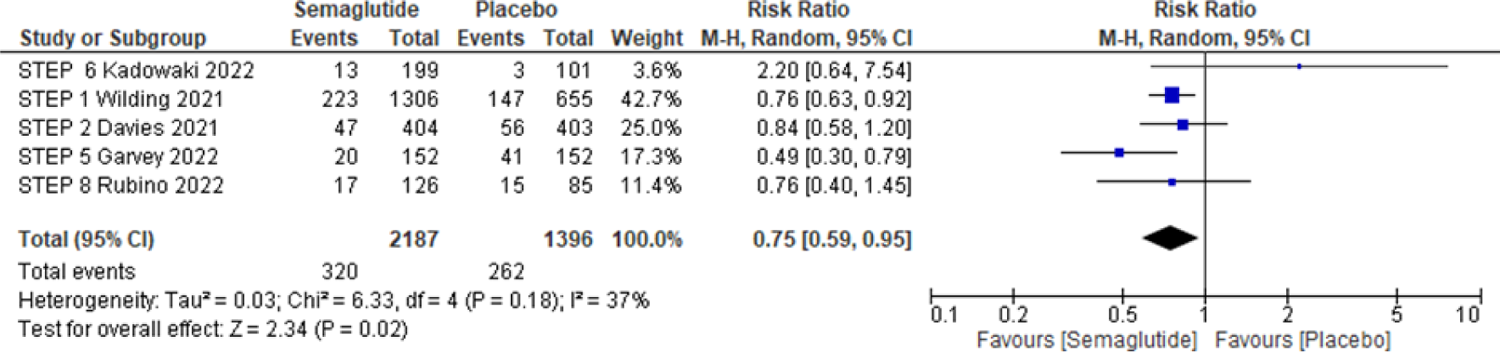
Forest plot for meta-analysis of the effect of semaglutide versus placebo on the number of dropouts at 68 weeks follow-up

**Figure S6.**
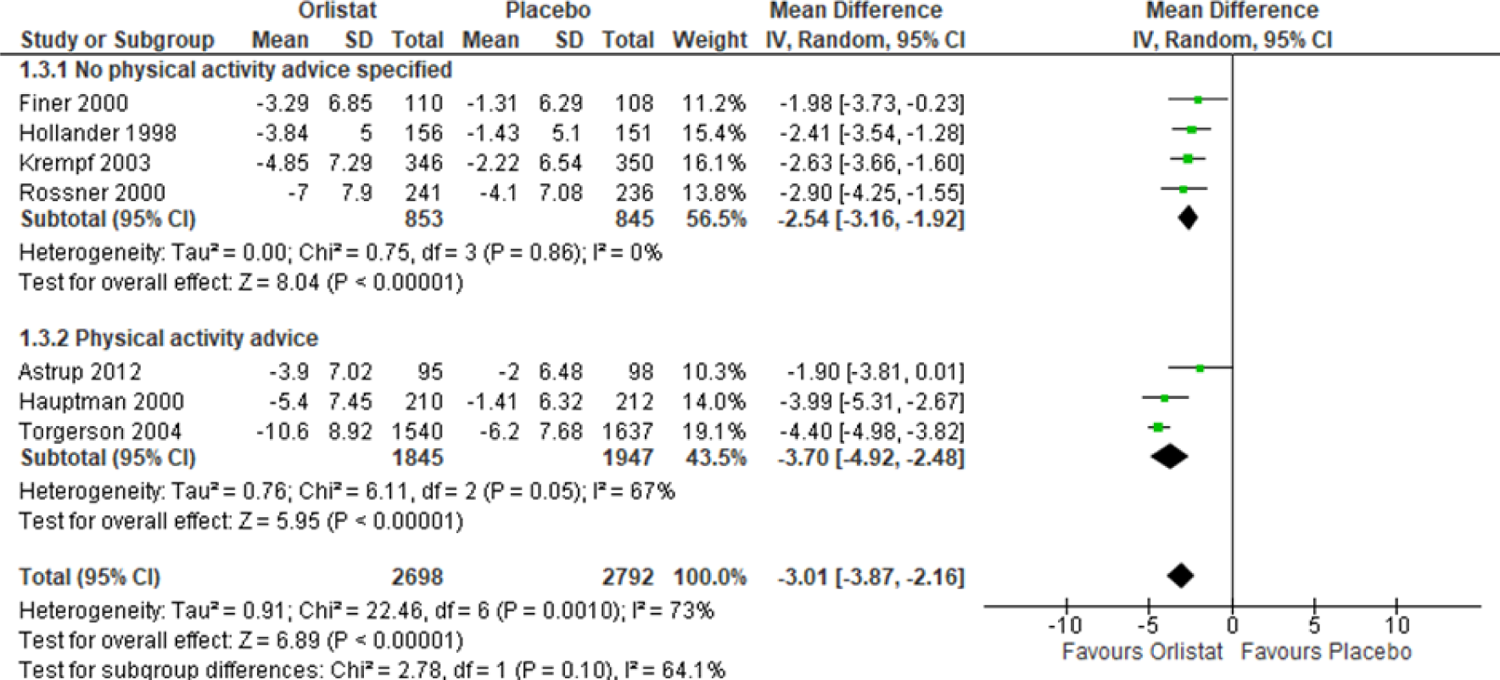
Last observation carried forward analysis of weight change in kg for orlistat trials

---

**Table S1.** Description of drug trials.

| Study ID<br>Location | Inclusion criteria | Run-in period | Intervention | No of contacts | Advisor |
| --- | --- | --- | --- | --- | --- |
| Astrup 2012<br>Location: 19 clinical research sites in 8 European countries | **Inclusion criteria:** men and women aged 18–65 years, of stable weight, with body mass index 30–40 kg m<sup>-2</sup> and fasting plasma glucose (FPG) of <7 mmol l<sup>-1</sup> (126 mg dl<sup>-1</sup>) at run-in.<br>**Exclusion criteria:** type 1 or 2 diabetes mellitus, drug-induced obesity, use of weight-lowering pharmacotherapy or participation in a weight-control study within the preceding 3 months, surgical obesity treatment and major medical conditions. There was no exclusion criterion based on psychiatric illness. | 2 weeks single-blind, placebo run-in. | **Diet:** 500 kcal/day deficit diet. About 30% of energy from fat, 20% from protein, and 50% from carbohydrates.<br>**Physical activity:** increased physical activity using pedometers.<br>**Resources provided:** 3-day food diary.<br>a. Liraglutide 3.0mg<br>b. Orlistat 120mg three times daily<br>c. Placebo | 20 in first year - weekly during the dose escalation period, 2 weekly in first 16 weeks of constant dose, otherwise every 4 weeks. | Unclear, possibly dietician. |
| Bakris 2002<br>Location: 41 referral centres in the continental USA | **Inclusion criteria:** ≥40 years, BMI 28-43kg/m<sup>2</sup>; ≥1 antihypertensive medication stable for at least 12 weeks preceding study entry; sitting DBP 96-109mmHg on 2 consecutive visits.<br>**Exclusion criteria:** unstable medical and/or psychiatric illness, initiation or change in diuretic therapy (within 12 weeks), previous weight reduction surgery, active GI disorders (except mild lactose intolerance/ diarrhoea/ constipation), history of bulimia, laxative abuse, substance abuse, alcohol abuse. Unwillingness/ inability to comply with protocol requirements; pregnant or lactating women, women of childbearing potential not on acceptable contraceptive methods; use of nicotine replacement therapy, appetite suppressants, fish-oil supplements, oral retinoids, chronic systemic steroids other than sex hormone replacement and GnRH, acute antidepressant or anxiolytic therapy. | Not described. | **Diet:** 600kcal/day deficit; ≥30% of calories as fat.<br>**Physical activity:** Encouraged to participate in moderate physical activity.<br>**Resources provided:** Literature made available to all participants.<br>a. Orlistat 120mg three times daily with meals<br>b. Placebo three times daily with meals | 12 | Unclear, possibly healthcare professional. |
| Berne 2005<br>**Location:** 16 primary care centres and six hospital-based diabetes clinics in Sweden | **Inclusion criteria:** Either sex, 30-75 years, BMI 28-40kg/m<sup>2</sup>, type 2 diabetics receiving treatment with either metformin alone or metformin + sulphonylurea; HbA1c 6.5-10%.<br>**Exclusion criteria:** treatment with insulin, recent MI, other significant peripheral vascular, cardiac, respiratory, renal, neurological, gastrointestinal or endocrine diseases or signs of fat-soluble vitamin deficiencies; participants taking medications influencing appetite, resins, fish oil supplements and retinoids. | 2 weeks of low-fat diet. | **Diet:** 600kcal/day deficit; 30% calories from fat.<br>**Physical activity:** Encouraged through 30-minute walk daily.<br>**Resources provided:** Dietary counselling included leaflets and food diary.<br>a. Orlistat 120mg three times daily with food<br>b. Placebo three times daily with food | 12 | Nurse or dietician. |
| Broom 2002<br>**Location:** 54 GP surgeries and 12 hospital clinics in the UK. | **Inclusion criteria:** 18-80 years, BMI ≥ 28kg/m<sup>2</sup>, at least one of: impaired glucose tolerance (serum glucose ≥ 8.0mmol/L, 2 hours after standard OGTT); hypercholesterolaemia (total serum cholesterol ≥ 5.2mmol/L or LDL ≥ 4.2mmol/L at screening); hypertension (sitting DBP 90-105mmHg); compliance 60% or more throughout the study.<br>**Exclusion criteria:** Pregnancy, lactation, women of childbearing potential not using adequate contraception, MI/CABG/PCI in prior 3 months; surgery for weight reduction, active GI disorders, history of post-surgical adhesions, excessive alcohol intake or substance abuse drugs which might alter body weight or plasma lipids such as appetite suppressants, lipid-lowering resins, retinoids and fish oil supplements; | 2 weeks single-blind placebo and trial diet. | **Diet:** 600kcal/day deficit (min 1200kcal/day); 30% calories from fats. Till month 6 then reduced a further 300kcal/day to week 52.<br>**Physical activity:** No details provided.<br>**Resources provided:** Food and beverage intake diary.<br>a. Orlistat 120mg three times daily with main meals<br>b. Placebo three times daily with main meals | 13 | Unclear, probably healthcare professional – 80% in GP centres rest in specialist obesity centres. |
|  | administration of systemic steroids (other than HRT) concomitant pharmacotherapy for type 2 diabetes, hypertension or hypercholesterolaemia. |  |  |  |  |
| Davidson 1999<br>**Location:** 18 US research centres | **Inclusion criteria:** Either sex, >18 years, BMI 30-43kg/m<sup>2</sup>, adequate contraception in women of childbearing potential.<br>**Exclusion criteria:** weight loss >4 kg in previous 3 months, frequently changed smoking habits or had stopped smoking in last 6 months, history of or presence of substance abuse, excessive alcohol intake, significant cardiac, renal, hepatic, gastrointestinal, psychiatric or endocrine disorder; drug treated type 2 diabetes mellitus, concomitant use of medications altering appetite or lipid levels. | 4 weeks of single-blind placebo run-in and diet, needed 75% or more treatment compliance by capsule count to be randomised. | **Diet:** 500-800kcal/day deficit; 30% energy intake from fats (If still losing weight in last 3 months of year 1 then energy intake increased 200-300kcal/day).<br>**Physical activity:** Encouraged by walking briskly 20-30 minutes 3-5 times per week.<br>**Resources available:** Food diaries kept by participants and used for counselling; 4 behaviour modification sessions on weight loss in year 1 then 4 weight maintenance seminars in year 2.<br>a. Orlistat 120mg three times daily for year 1<br>b. Placebo three times daily for year 1 and year 2<br>rerandomized at weeks 52 to:<br>c. Placebo 3 times daily<br>d. Orlistat 120mg 3 times daily<br>e. Orlistat 60mg 3 times daily | 18 | Dietician. |
| Finer 2000<br>**Location:** 5 centres in the UK | **Inclusion criteria:** Either sex; 18 years or more, BMI 30-43 kg/m<sup>2</sup>, women of childbearing potential if using adequate contraceptive precautions, >75% compliance (returned tablets) during run-in phase.<br>**Exclusion criteria:** weight loss >4kg in the 3 | 4 week single-blind placebo with diet, needed >75% tablet compliance to | **Diet:** 600kcal/day deficit (lowest intake 1200kcal/day); 30% from fat. Alcohol consumption limited to 150g/week. Prescribed daily energy intake was further reduced by approximately 300kcal/day in | 17 | Not stated. |
|  | months before screening; history of any serious systemic disease, including diabetes, uncontrolled hypertension, previous surgery for weight reduction, history of post-surgical adhesions, history/presence of cancer, psychiatric or neurological disorder requiring chronic medications or liable to prejudice patient compliance; evidence of alcohol or substance abuse; bulimia or evidence of laxative abuse; pregnancy or lactation; post-menopausal women who had been amenorrhoeic for less than 1 year. Patients were excluded if they had taken drug capable of influencing body weight, resins for lipid lowering, anti-coagulants, digoxin or lipid-soluble vitamin supplements within the previous month. | enter main study. | all patients at the end of week 24, regardless of whether body weight had stabilized.<br>**Physical activity:** No details provided.<br>**Resources provided:** 4-day food diaries used at intervals for individual dietician advice.<br>a. Orlistat 120mg three times daily with meals<br>b. Placebo orally three times daily with meals |  |  |
| Hauptman 2000<br>**Location:** 17 primary care centres in the US | **Inclusion criteria:** Either sex, >18 years, BMI 30-44 kg/m<sup>2</sup>, completed 4-week pre-treatment phase with 75% or more compliance (by capsule count).<br>**Exclusion criteria:** pregnancy, lactation, women of childbearing potential not taking adequate contraception; weight loss >4kg last 3 months, history of significant cardiac, renal, hepatic, gastrointestinal disorders; uncontrolled hypertension or other clinically significant condition; gastrointestinal surgery for weight reduction; bulimia or laxative and/or substance abuse, abnormal laboratory measures (values 10% or greater than reference value for the normal range and sufficient to require medical follow-up by study physician); change in smoking habits in previous 6 months, use of any drug | 4 weeks single blind placebo and diet, needed 75% tablet compliance for randomisation. | **Diet:** 1200kcal/day (weight <90kg initially) or 1500kcal/day (weight ≥90kg initially); 30% from fats, 50% CHO, 20% protein, maximum 300mg/day cholesterol, maximum 10 drinks/week alcohol; dietary guidance on intake from study physician at start of pre-treatment only.<br>Intake increased by 300kcal/day for participants still losing weight at end week 52 or no dietary adjustment for those weight stable until week 104.<br>**Physical activity:** encouraged by brisk walking 20-30 minutes 3-5 times/week. | 18 | Physician (dietitians and behavioural psychologists not involved at any stage of the trial). |
|  | which might influence body weight or food intake in 8 weeks prior to screening. |  | **Resources provided:** Dietary records kept 10 times during study; Videos on behaviour modification techniques for weight control 4 times in first 52 weeks, pamphlets for weight maintenance given 4 times during weeks 53-104 based on "Live for Life" program.<br>a. Orlistat 120mg three times daily with main meals<br>b. Orlistat 60mg three times daily with main meals<br>c. Placebo three times daily with main meals |  |  |
| Hollander 1998<br>**Location:** 12 diabetic clinic centres in USA | **Inclusion criteria:** Either sex, >18yrs, BMI 28-40 kg/m<sup>2</sup>, type 2 diabetes, clinically stable on glyburide or glipizide for 6 months or more; HbA1c% 6.5-10% at screening, fasting plasma glucose 5.6-12.2mmol/L at end of 4th week of pre-treatment, blood levels of fat-soluble vitamins above lower limit of normal reference range, completion and compliance by tablet count of 70% or more during pre-treatment.<br>**Exclusion criteria:** pregnancy, lactation, women of childbearing potential not taking adequate contraceptive measures, clinically relevant conditions e.g. psychiatric disorders, substance abuse, cholecystitis, pancreatic disease, uncontrolled hypertension; significant complications associated with diabetes, weight loss of >4kg during last 3 months, history of recurrent nephrolithiasis or symptomatic<br>cholelithiasis; gastrointestinal surgery for weight reduction, history of bulimia or laxative abuse or if they had taken any drug that might influence body weight or plasma lipids in 8 weeks prior to start of study. | 5 week single-blind placebo and study diet, needed at least 70% compliance to be randomised. | **Diet:** mildly hypocaloric diet during run-in, then 500kcal/day deficit from baseline to week 52.<br>**Physical activity:** No details provided.<br>**Resources provided:** Additional diet counselling and a standardised commercially available vitamin supplement given if 2 consecutive vitamin measures fell below reference range.<br>a. Orlistat 120mg three times daily taken with meals.<br>b. Placebo three times daily taken with meals. | Unclear between 16-18. | Not stated. |
| Kelley 2002<br>**Location:** 43 centres in the US | **Inclusion criteria:** Men and women, 40-65 years, BMI 28-43 kg/m<sup>2</sup> and a stable weight (< 3 kg weight change) for 3 months before study entry, treatment with a stable daily dose of insulin for 6 weeks before study entry and HbA1c 7.5-12.0% at screening, women with childbearing potential were required to have a negative pregnancy test and to use an acceptable form of contraception throughout the study.<br>**Exclusion criteria:** Treatment with thiazolidinedione for diabetes, change in diabetes medication in last 12 months (except insulin), medical history or presence of renal, hepatic or endocrine disorders, previous bariatric surgery, use of approved/ experimental weight reduction medications or treatments, presence of malabsorption syndrome, bulimia or laxative abuse, disorders that could affect compliance with the requirements of the study. | Not described. | **Diet:** Nutritionally balanced diet designed for 0.25-0.5 kg loss every week. 30% of calories from fat, 50% carbohydrate, 20% protein and max. 300 mg/day cholesterol. Prescribed caloric intake reduced by 200 kcal/day (with min. intake 1200 kcal/day) at week 24.<br>**Physical activity:** Encouraged to participate in moderate physical activity. Withdrawn if compliance <60%<br>**Resources provided:** Lifestyle and behavioural modification literature were available to all patients. Dietary compliance was monitored by the use of dietary intake records.<br>a. Orlistat 120 mg three times daily.<br>b. Placebo three times daily. | 13 | Initially a registered dietician. |
| Krempf 2003<br>**Location:** 81 hospital centres in France | **Inclusion criteria:** Men and women aged 18-65, BMI ≥ 28kg/m<sup>2</sup>.<br>**Exclusion criteria:** serious eating disorders or type 1 or type 2 diabetes, pregnant or lactating women; smoking 1 or more packs of cigarettes per day or intention to stop smoking during the | 15-day placebo run-in. | **Diet:** 20% energy reduction (initial intake assessed through food diaries). Fat intake 30%. The diet energy prescription was decreased by a further 10% (never below 1200kcal/day) in patients with | 13 based on Fig 1. | Dietitian, possibly other healthcare staff. |
|  | trial; previous surgical treatment for obesity; known or suspected substance abuse; significant thyroid, renal, hepatic, gastrointestinal or immune disorders, or concomitant use of medications that alter body weight, appetite or the absorption of food. |  | stable weight (+/- 0.5kg since last visit) or weight gain.<br>**Physical activity:** No details provided.<br>**Resources provided:** Food diaries used to assess understanding during run-in period.<br>a. Orlistat 120mg three times daily.<br>b. Placebo three times daily for 18 months. |  |  |
| Lindgarde 2000<br>**Location:** 33 primary care centres in Sweden | **Inclusion criteria:** men and non-pregnant women, 18-75 years, BMI 28-38kg/m<sup>2</sup>, fasting serum glucose $\geq 6.7$ mmol/L/confirmed type 2 diabetes treated with sulfonylurea/ metformin; total serum cholesterol $\geq 6.5$ mmol/L and/or LDL cholesterol $\geq 4.2$ mmol/L on at least 2 occasions or prescribed lipid lowering medications; DBP $\geq 99$ mmHg on at least 2 occasions or confirmed hypertensive treated with antihypertensive medication.<br>**Exclusion criteria:** insulin treated participants, women of child-bearing potential who were lactating or using inadequate contraception; MI within 3 months prior to screening, surgery for weight reduction, active GI disorders (with the exception of controlled lactose intolerance), pancreatic disease, history of post-surgical adhesions, excessive alcohol or substance abuse, participants requiring any drug which might alter body weight or plasma lipids, such as appetite suppressants, lipid-lowering resins, retinoids or fish oil supplements; systemic steroids (other than HRT and insulin). | 2 weeks single blind placebo plus mildly hypocaloric diet. | **Diet:** 600kcal/day deficit (minimum 1200kcal/day); 30% energy from fat continued up to month 6 when energy content reduced additional 300kcal/day.<br>**Physical activity:** Encouraged by taking 30 minutes' walk each day.<br>**Resources provided:** Participants also received dietary counselling as part of self-help weight control educational package including leaflets and videotape given at start of run-in phase.<br>a. Orlistat 120mg three times daily<br>b. Placebo three times daily | Contacted 11 times (baseline, twice in first month then monthly to month 6 then every 2 months to month 12). | Practice nurse. |
| Miles 2002<br>**Location:** 34 centres in the US and 6 centres in Canada | **Inclusion criteria:** Patients with type 2 diabetes who were 40-65 years of age, BMI 28-43kg/m<sup>2</sup>, had maintained a stable weight for 2-3 months, HbA1c 7.5-12.0% and received metformin treatment at 1,000-2,550mg/day for at least 6 weeks. Sulfonylurea therapy in combination with metformin was permitted as long as the sulfonylurea dose was stable for 12 weeks before study entry.<br>**Exclusion criteria:** Patients receiving insulin, thiazolidinediones, or a-glucosidase inhibitors; any clinical condition that might affect study end points, including renal, hepatic, or endocrine disorders, poorly controlled hypertension (SBP ≥160mmHg or DBP ≥100mmHg) active gastrointestinal disease, previous bariatric surgery, a history of bulimia, substance abuse, or the use of any weight loss medications. Women who were pregnant, lactating, or of childbearing potential were also excluded. | Unclear, 2-week screening phase. | **Diet:** 600kcal/day deficit; 30% as fat, 50% as CHO and 20% as protein, with a maximum cholesterol content of 300mg/day. Intake was reduced by an additional 200kcal after 6 months.<br>**Physical activity:** 'encouraged to increase their level of physical activity'.<br>**Resources provided:** Dietary counselling at baseline and at regular intervals throughout the study.<br>a. Orlistat 120mg three times daily with main meals.<br>b. Placebo three times daily with main meals. | 13 | Unclear, possibly healthcare professional. |
| Rossner 2000<br>**Location:** 14 European centres | **Inclusion criteria:** Either sex, 18 years or more, BMI 28-43kg/m<sup>2</sup>, completed 4-week pre-treatment phase and 75% or more compliance (by capsule count).<br>**Exclusion criteria:** pregnancy, lactation, women of childbearing potential not taking adequate contraception, clinically significant conditions (excluding obesity) that might affect study outcome, >4kg weight loss in previous 6 months, | 4-week single blind placebo run-in and at least 75% drug compliance needed for randomisation. | **Diet:** 600kcal/day deficit, 30% from fat. 2nd year diet was adjusted; lost ≥3kg between weeks 40-52 intake was prescribed at estimated energy intake minus 10% kcal/day; lost <3kg no dietary adjustment made.<br>**Physical activity:** No details | 13; contacted baseline, every 2 weeks for first 2 months then monthly up to month 6 then every 2 months to | Dietitian, possibly other healthcare staff. |
|  | gastrointestinal surgery for weight loss, history of post-surgical adhesions or of bulimia or laxative abuse; any drug that might influence body weight or serum lipids taken in 8 weeks prior to screening; uncontrolled hypertension, drug treated diabetes mellitus, history or presence of symptomatic cholelithiasis. |  | provided.<br>**Resources provided:** 4-day food diaries.<br>a. Orlistat 60mg three times daily.<br>b. Orlistat 120mg three times daily.<br>c. Placebo 3 times daily. | month 24. |  |
| Sjostrom 1998<br>**Location:** 15 European centres | **Inclusion criteria:** Either sex, 18 years or more, BMI 28-47kg/m<sup>2</sup>, women of childbearing potential if using adequate contraception, >75% compliance during pre-treatment phase at end of year 1 in order to continue to year 2.<br>**Exclusion criteria:** serious diseases including uncontrolled hypertension (DBP 105mmHg or more) and pharmacologically treated diabetics, weight loss >4kg in 3 months prior to screening, surgery for weight reduction, history of post-surgical adhesions, bulimia or laxative abuse, use of any drug which might influence body weight or plasma lipids in last month; drug or alcohol abuse. | 4 week single-blind run-in with >75% compliance required for run-in. | **Diet:** 600kcal/day deficit (minimum 1200kcal/day); 30% from fats for run-in and first 24 weeks; then until week 52 reduced additional 300kcal/day (minimum 1000kcal/day); 30% fats, 50% CHO, 20% protein, 300mg/day cholesterol, 3 main meals and optional snack daily, 150mg/week alcohol.<br>Year 2 maintenance diet.<br>**Physical activity:** No details provided.<br>**Resources provided:** No details provided.<br>a. Orlistat 120mg three times daily.<br>b. Placebo three times daily. | 17 in first year. | Dietician. |
| Swinburn 2005<br>**Location:** Eight clinical | **Inclusion criteria:** Men and women, aged 40–70, with a BMI between 30 and 50 kg/m<sup>2</sup> and one of the following: hypercholesterolemia (serum total cholesterol of >5.5 mmol/l and/or LDL-cholesterol of >3.5 mmol/l and clinically stable if | 4-week placebo run-in with dietary and physical activity advice. | **Diet:** Reduce daily dietary fat intake to 25-30% or about 40 g/day.<br>**Physical activity:** Advice given about undertaking regular, | 15 | Dietician. |
| research centres in Australia and New Zealand | on treatment); or hypertension (systolic >140mmHg and/or diastolic >90mmHg and clinically stable if on treatment); or type 2 diabetes treated with dietary modification or any oral hypoglycaemic agent for at least 6 months and clinically stable (glycated haemoglobin: 6.5–10%).<br>**Exclusion criteria:** A history of significant cardiac, renal, hepatic, gastrointestinal or endocrine disorders; uncontrolled hypertension; previous gastrointestinal surgery for weight reduction; a history of post-surgical adhesions; smoking; a history or presence of substance abuse, bulimia, type-1 diabetes, psychiatric disorders or active gastrointestinal disease. |  | moderate-intensity physical activity of at least 30 min/day on most days.<br>**Resources provided:** Advice from a dietician about identifying the sources of dietary fat (such as through label reading) and reducing them as much as possible.<br>a. Orlistat 120mg three times daily.<br>b. Placebo tablet three times daily. |  |  |
| Torgerson 2004<br>**Location:** 22 Swedish medical centres | **Inclusion criteria:** 30–60 years old and with at least BMI 30.0 kg/m<sup>2</sup>. The stipulated sex distribution was 55% women and 45% men. Also, subjects had to be nondiabetic as determined by a 2-hour oral glucose tolerance test (OGTT) at the baseline examination. However, at least 10% of the subjects were requested to have impaired glucose tolerance (IGT).<br>**Exclusion criteria:** A change in body weight > 2 kg between the screening and baseline examinations and a blood pressure above 165 mm Hg systolic or 105 mm Hg diastolic on the same two consecutive visits. Diabetes mellitus, myocardial infarction within 6 months, symptomatic cholelithiasis, gastrointestinal surgery for weight reduction or peptic ulcer, active gastrointestinal disorder or pancreatic<br>disease, malignancy, significant psychiatric or neurologic disorder, abuse or previous participation in any trial of orlistat were not allowed. | Not described. | **Diet:** ~800 kcal/day deficit, 30% fat and <300 mg of cholesterol per day. The prescribed energy intake was readjusted every 6 months.<br>**Physical activity:** encouraged to walk at least 1 extra kilometre a day in addition to their usual physical activity.<br>**Resources provided:** Dietary counselling every 2 weeks for the first 6 months and monthly thereafter.<br>a. Orlistat 120 mg three times daily.<br>b. Placebo three times daily. | At least 20. | Unclear, probably healthcare professional. |
| Davies 2015<br>**Location:**<br>126 sites in 9 countries (France, Germany, Israel, South Africa, Spain, Sweden, Turkey., United Kingdom, United States) | **Inclusion criteria:** ≥18 years, BMI ≥27, with a stable body weight (<5kg in last 3 months), diagnosed T2DM treated with diet and exercise alone or in combination with 1 to 3 oral hypoglycaemic agents.<br>**Exclusion criteria:** eTable 1 – treatment with GLP-1 receptor agonists, DPP-4 inhibitors or insulin in the last 3 months; recurrent hypoglycaemia or hypoglycaemic unawareness; proliferative retinopathy pr maculopathy; untreated or uncontrolled hypothyroidism/hyperthyroidism; chronic or idiopathic acute pancreatitis; obesity caused by endocrine disorder; medications that may cause significant weight gain; diet attempts using herbal supplements or OTC medications in 3 months prior to screening; surgical treatment for obesity; family or personal history of MEN2 Or familial MTC; major psychiatric history; uncontrolled hypertension; females who may become pregnant; LANGUAGE BARRIER etc. | Not described. | **Diet:** Participants were encouraged to follow a diet containing a maximum of 30% of energy from fat, approximately 20% of energy from protein, and approximately 50% of energy from carbohydrates, with a 500-kcal/d deficit based on estimated total energy expenditure.<br>**Physical activity:** exercise program (≥150 min/week of brisk walking).<br>**Resources provided:** pedometer and 3-day food diary.<br>a. Liraglutide 3.0mg – starting dose of 0.6mg escalated in increments of 0.6mg weekly to the treatment dose.<br>b. Liraglutide 1.8mg - as above.<br>c. Placebo.<br>Participants who discontinued were asked to return at week 56 for follow-up. | 21 - The trial consists of a screening visit, a randomisation visit, 15 treatment visits and 4 follow-up visits. | Dietician. |
| Garvey 2020 | **Inclusion criteria:** ≥18 years with a BMI ≥27, stable body weight, diagnosed with T2DM with a Hba1c 6-10% at screening and receiving stable treatment with any basal insulin and ≤2 OADs.<br>**Exclusion criteria:** T1DM, recurrent severe hypoglycaemic episodes within the last year, or | Not described. | **Intensive behavioural therapy**<br>**Diet:** 1200kcal/day for individuals <200lb, the sum of weight (lb)x6 (kcal/lb) in individuals 200-300lbs and 1800kcal/day for those >300lbs | Individuals attended a total of 23 individual or group counselling | Delivered by a registered dietitian or similarly qualified health care |
|  | use of DP4 inhibitors, GLP-1 receptor agonists, bolus insulin or medications known to induce significant weight change in the previous 90 days. History of cardiovascular event, medullary thyroid carcinoma or MEN type 2; pregnancy; breastfeeding; or intention to become pregnant; history of pancreatitis. |  | **Physical activity:** increased physical activity of moderate intensity, starting at 100 min/week, and increasing by 25 minutes every 4 weeks to a recommended 250min/week.<br>**Resources provided:** behavioural therapy delivered in frequent 15-minute brief counselling sessions.<br>a. Liraglutide 3.0mg – starting at 0.6mg then increasing by 0.6mg weekly.<br>b. Placebo. | sessions during the 56-week period. | professional. |
| O’Neil 2018<br>**Location:** 8 countries involving 71 clinical sites: Australia (n=5), Belgium (n=5), Canada (n=9), Germany (n=6), Israel (n=7), Russia (n=10), the UK (n=8), and the USA (n=21) | **Inclusion criteria:** 18 years or older without diabetes, and with a body-mass index (BMI) of 30 kg/m<sup>2</sup> or more that was not of endocrine aetiology (e.g., Cushing’s syndrome). Self-reported bodyweight must not have fluctuated by more than 5 kg in the 90 days before screening. Eligible individuals must have undergone at least one previous unsuccessful nonsurgical weight-loss attempt and been free from major depressive symptoms (defined as a screening Patient Health Questionnaire-9 [PHQ-9] score. | 1 week screening period. | **Diet:** Participants were advised to follow a daily energy intake limit of approximately 500 kcal below their total energy expenditure, estimated from their basal metabolic rate using a method described elsewhere. A maintenance diet without an energy deficit was recommended to participants if their BMI declined to 22 kg/m<sup>2</sup> or less.<br>**Physical activity:** physical activity level of 1·3. Physical activity counselling was based on participant capability, emphasising a recommended minimum activity time of 150 min per week without specifying exercise intensity.<br>**Resources provided:** Nutritional<br>compliance was assessed, and nutritional and physical activity counselling was provided every 4 weeks. Compliance was assessed on a 10-point numeric rating scale from 0 (not at all compliant) to 10 (fully compliant) monthly from week 4.<br>a. Liraglutide 3.0mg.<br>b. Placebo. | 21 - screening, baseline, every 2 weeks through week 20, and every 4 weeks to week 52, plus follow-up at week 59. | Qualified research staff. |
| Pi-Sunyer 2015<br>**Location:** 191 sites in 27 countries in Europe, North America, South America, Asia, Africa and Australia. | **Inclusion criteria:** ≥18 years with a stable body weight, either a BMI ≥30 or ≥27 + treated or untreated dyslipidaemia or hypertension.<br>**Exclusion criteria:** type 1 or 2 diabetes, use of medications that cause clinically significant weight gain or loss, previous bariatric surgery, history of pancreatitis, history of major depressive disorder or other severe psychiatric disorders, a family or personal history of multiple endocrine neoplasia type 2 or familial medullary thyroid carcinoma. | Not described. | **Diet:** 500kcal/day deficit with 30% energy from fat, 20% from protein and 50% from carbohydrates.<br>**Physical activity:** increased to at least 150 minutes per week.<br>**Resources provided:** Counselling on lifestyle modification in individual or groups sessions. Pedometers and 3-day food diaries to encourage adherence.<br>a. Liraglutide 3.0mg – starting at 0.6mg then increasing by 0.6mg weekly.<br>b. Placebo.<br>Those randomised to Liraglutide were rerandomized at week 56 to continue liraglutide or switch to placebo for 12 weeks. | 17 – “All patients received standardized counselling on lifestyle modification approximately monthly” | Dietician. |
| Rubino 2022 STEP 8 | **Inclusion criteria:** ≥18 years with 1 or more self-reported unsuccessful dietary efforts to lose | Not described. | **Diet:** 500kcal per day reduction relative to the estimated total | Counselling every 4-6 | Qualified healthcare |
| | weight and either a BMI $\geq 30$ or $\geq 27 + 1$ or more treated or untreated weight-related condition.<br>**Exclusion criteria:** diabetes, HbA1c $\geq 48$, self-reported weight change $> 5$ kg within 90 days before screening. | | energy expenditure.<br>**Physical activity:** 150 minutes per week.<br>**Resources provided:** no details provided.<br>a. Semaglutide 2.4 mg – started at 0.25 mg per weeks and escalated in a fixed-dose regimen every 4 weeks until the target dose was reached.<br>b. Liraglutide 3.0 mg – started at 0.6 mg and escalated to 3.0 mg over 4 weeks.<br>c. Matching placebos. | weeks via visits or telephone contact. | professional. |
| Wadden 2020 | **Inclusion criteria:** $\geq 18$ years with a BMI $\geq 30$, stable body weight.<br>**Exclusion criteria:** HbA1c $\geq 6.5\%$, type 1 or type 2 diabetes, use of medications known to induce significant weight loss or gain, inadequately treated hypertension, pregnancy or breastfeeding, history of CVD, severe congestive heart failure, second degree or greater heart block, medullary thyroid carcinoma, MEN type 2, pancreatitis, major depressive disorder within the past 2 years, history of suicide attempt or malignancy within the past 5 years. | Not described. | **Diet:** Participants who weighed $< 91$ kg ( $< 200$ lb) at randomisation were prescribed 1200 kcal/day; the caloric prescription for those who weighed 91-136 kg (200-300 lb) was calculated by body weight (pounds) $\times 6$ (kilocalories per pound), and participants who weighed $> 136$ kg ( $> 300$ lb) were prescribed 1,800 kcal/d. Diet recommendations including approximately 15% to 20% of kilocalories from protein, 20% to 35% from fat, and the remainder from carbohydrates.<br>**Physical activity:** initially prescribed 100 min/week of moderate-intensity physical<br>activity (e.g., brisk walking) encouraged in bouts of 10 minutes or more and spread their activity across 4 to 5 days each week. Physical activity was increased by 25 minutes every 4 weeks, with a goal of 250 min/week.<br>**Resources provided:** behavioural therapy delivered in frequent 15-minute brief counselling sessions. Participants were encouraged to attend the counselling visits regardless of whether they discontinued study medication.<br>A. Liraglutide 3.0mg – starting at 0.6mg then increasing by 0.6mg weekly.<br>B. Placebo | 23 - “attended a mean of 22.5 and 21.2 visits, respectively, of 23 possible treatment visits”. | Registered dietitians - attended an in-person 2.5-hour training that reviewed key IBT principles and practices, as presented in the provider and participant treatment manuals used in this study. |
| Davies 2021 STEP 2<br>**Location:** 149 outpatient clinics in 12 countries across Europe, North America, South | **Inclusion criteria:** ≥18 years with 1 or more self-reported unsuccessful dietary efforts to lose weight and either a BMI ≥27, Hba1c 7-10% and diagnosed with T2DM at least 180 days before screening.<br>**Exclusion criteria:** changes in body weight >5kg in 90 days before screening, planned or previous surgical obesity treatment or weight loss device. | Not described. | **Diet:** 500kcal per day reduction relative to the estimated total energy expenditure calculated at the time of random allocation.<br>**Physical activity:** 150min per week – e.g., walking or using the stairs.<br>**Resources provided:** Patients were instructed how to measure their physical activity and food intake and were encouraged to keep a food diary and activity diary daily, which was reviewed during counselling sessions. | 17 - Every 4<sup>th</sup> week, via in-person visit or telephone. | Dietician or a similarly qualified healthcare professional. |
| America, the Middle East, South Africa, and Asia |  |  | a. Semaglutide 2.4mg – started at 0.25mg per weeks and escalated in a fixed-dose regimen every 4 weeks until the target dose was reached.<br>b. Semaglutide 1.0mg – as above.<br>c. Placebo.<br>Patients remained in the trial regardless of whether they discontinued treatment using the study drug |  |  |
| Garvey 2022 STEP 5<br>**Location:** 41 sites across 5 countries (Canada, Italy, Hungary, Spain and the United States) | **Inclusion criteria:** ≥18 years with 1 or more self-reported unsuccessful dietary efforts to lose weight and either a BMI≥30 or ≥27+1 or more treated or untreated weight-related condition.<br>**Exclusion criteria:** history of type 1 or type 2 diabetes; self-reported weight change >5kg within 90 days before screening; previous or planned obesity surgery or weight loss device; treatment with any anti-obesity medication in 90 days before screening; uncontrolled thyroid disease; major psychiatric history. | Not described. | **Diet:** 500kcal/day deficit diet.<br>**Physical activity:** 150min/week physical activity.<br>**Resources provided:** recording of diet and physical activity via a diary, app or other tools.<br>a. Semaglutide 2.4mg – started at 0.25mg per weeks and escalated in a fixed-dose regimen every 4 weeks until the target dose was reached at week 16.<br>b. Placebo. | 30 - Every 2<sup>nd</sup> week till week 16 then every 4<sup>th</sup> week till week 104 then at 7 weeks follow-up. | Dietician or similarly qualified healthcare professional. |
| Kadowaki 2022 STEP 6<br>**Location:** 22 sites in Japan | **Inclusion criteria:** ≥18 years in South Korea; ≥20 years in Japan, at least one self-reported unsuccessful dietary attempt to lose bodyweight, BMI ≥27.0 kg/m<sup>2</sup> + 2 or more treated or untreated weight-related | Not described. | **Diet:** 500kcal per day reduction relative to the estimated total energy expenditure calculated at the time of random allocation.<br>**Physical activity:** 150min per week | 17 - Counselling every 4<sup>th</sup> week via visits or telephone | Dietician or similar qualified healthcare professional. |
| and 6 sites in South Korea | comorbidities, or BMI $\geq 35.0 \text{ kg/m}^2 + 1$ or more treated or untreated weight-related comorbidity according to the JASSO guidelines. At least one comorbidity had to be hypertension or dyslipidaemia, or, in Japan only, type 2 diabetes. **Exclusion criteria:** self-reported weight change $>5\text{kg}$ within 90 days before screening, planned or previous surgical obesity treatment or medication. | | – e.g., walking or using the stairs. **Resources provided:** Instruction on how to measure food intake and physical exercise were provided and participants were encouraged to record these measurements daily using paper, an app or similar tool.<br>a. Semaglutide 2.4mg – started at 0.25mg per weeks and escalated in a fixed-dose regimen every 4 weeks until the target dose was reached.<br>b. Semaglutide 1.7mg – as above.<br>c. Placebo. | contact. | |
| Wilding 2021 STEP 1<br>**Location:** 129 sites in 16 countries in Asia, Europe, North America, and South America | **Inclusion criteria:** $\geq 18$ years with 1 or more self-reported unsuccessful dietary efforts to lose weight and either a BMI $\geq 30$ or $\geq 27+1$ or more treated or untreated weight-related condition. **Exclusion criteria:** diabetes, HbA1c $\geq 48$, history of chronic pancreatitis, acute pancreatitis within 180 days before enrolment, previous surgical obesity treatment, use of AOM within 90 days of enrolment. | Not described. | **Diet:** reduced-calorie diet (500kcal deficit per day relative to energy expenditure at time of randomisation)<br>**Physical activity:** increased physical activity (150 minutes/week encouraged).<br>**Resources provided:** Both diet and activity were recorded daily in a diary or by use of a smartphone application or other tools and were reviewed during counselling sessions.<br>a. Semaglutide, administered with a prefilled pen injector, was initiated at a<br>dose of 0.25mg once weekly for the first 4 weeks, with dose increased every 4 weeks to reach the maintenance dose of 2.4mg weekly by week 16.<br>b. Placebo.<br>Participants discontinuing treatment prematurely remained in the trial. | 17 – every 4 weeks. | Dietician or a similar qualified healthcare professional. |

**Table S2.** Description of low-fat reducing diet trials.

| Study ID<br>(Citation)<br>**Location** | Inclusion criteria | Run-in<br>period | Intervention | Number of<br>contacts | Advisor |
| --- | --- | --- | --- | --- | --- |
| Abbenhardt<br>2013<br>Abbenhardt<br>et al., 2013<br>**Location:**<br>Greater<br>Seattle area<br>(WA, USA) | **Inclusion criteria:** Female, 50-75 years, BMI≥25kg/m<sup>2</sup> (if Asian-American ≥23kg/m<sup>2</sup>), <100min moderate activity per week; postmenopausal; fasting glucose<7mmol/L and not taking diabetic medications; non-smoking; alcohol intake<2 drinks per day; ability to attend diet or exercise sessions at the intervention site; normal exercise tolerance test<br>**Exclusion criteria:** menopausal hormone therapy for the past 3 months; history of breast cancer, heart disease, diabetes mellitus or other serious medical conditions | No<br>details<br>given | **Diet:** Goal of 1200-2000kcal/day based on weight, <30% fat.<br>**Physical activity:** 45 minutes of moderate-to-vigorous intensity exercise 5 days per week for 12 months. 3 supervised sessions/week at the facility and 2 days/week at home.<br>**Control:** Asked not to change their diet or exercise habits. After 12 months, women were offered four weight loss classes ad 8 weeks of facility exercise training.<br>a. Control<br>b. Aerobic exercise<br>c. Dietary weight loss<br>d. Diet and exercise | Weekly<br>dietician-led<br>group<br>meetings<br>months 1-6,<br>monthly<br>group<br>meetings +<br>phone/email<br>contact with<br>dietitians'<br>months 7-12.<br>At least 32<br>face-to-face<br>for diet<br>group<br>At least 2 for<br>control | Dieticians<br>with training<br>in behaviour<br>modification |
| HOT 1999<br>Jones et al.,<br>1999<br>**Location:**<br>University<br>of<br>Mississippi, | **Inclusion criteria:** Either sex, >50 years, BMI not less than 27kg/m<sup>2</sup>, baseline DBP >100mm Hg<br>**Exclusion criteria:** | No<br>details<br>given | **Diet:** Counselling on food selection and preparation and establishing weight reduction goals, calorie and fat restriction; counselled again at 2-4 weeks and attended group support sessions twice monthly for first 3 months then every 3-6 months, weight measured at 6 monthly intervals<br>**Control:** Told to try to lose weight but no formal counselling | a. 8 (based<br>on text and<br>graph) b. 3 | Registered<br>dietician in<br>diet group |
| USA |  |  | a. Diet<br>b. Control |  |  |
| Lim 2010<br>Lim et al., 2010<br>**Location:** Adelaide, Australia | **Inclusion criteria:** age 20-65, at least one CVD risk factor (blood pressure and serum lipids) other than obesity; BMI 28-40kg/m<sup>2</sup><br>**Exclusion criteria:** use of hypoglycaemic medication or drugs that affect insulin sensitivity, a history of heavy alcohol consumption, a history of metabolic or coronary heart disease, type 1 or type 2 diabetes, widely fluctuating exercise patterns and frequent dining out (>2 per week and unable to cease). | No details given | **Diet:** 12-week intensive phase of 8 weeks weight loss diet followed by energy intake titration upwards to restore the daily energy deficit. Then advised to maintain their allocated energy-restricted diet for an additional 12 months. Diets had the same energy content (1550kcal). Meal plans, recipes and specific quantities of key foods were provided at baseline. Subjects in the diet groups completed a 3-day weighed food diary (2 weekdays and 1 weekend day) at 3, 6, 9, 12, and 15 months, reviewed and analysed by a qualified dietitian using Diet/1 Nutrition Calculation<br>a. VLC diet (35% protein, 60% fat 20% saturated fat, 4% carbohydrate)<br>b. VLF (20% protein, 10% fat, 3 % saturated fat, 70% carbohydrate)<br>c. HUF (20% protein, 30% fat, 6% saturated fat, 8% polyunsaturated fat, 50% carbohydrate)<br>d. Control | 11 | Dietician |
| ODES 1995<br>Anderssen et al., 1995<br>**Location:** Ullevaal hospital, Oslo, Norway | **Inclusion criteria:** Either sex, 41-50 years, sedentary (exercise no more than once a week), BMI >24kg/m<sup>2</sup>, DBP 86-99mmHg, total cholesterol 5.2-7.74 mmol/l, HDL cholesterol <1.2 mmol/l, fasting serum triglycerides >1.4 mmol/l<br>**Exclusion criteria:** overt cardiovascular disease, diabetes, other disease or drugs that could interfere with the test results, treatment with anti-hypertensive drugs, | No details given | **Diet:** Counselling with spouse at baseline then individually at 3- and 9-months follow-up; diet adapted to individuals risk profile, main focus on energy restriction in those overweight, increase of fish products and vegetables, decrease of saturated fat, cholesterol and sugar, and salt restriction for participants with elevated BP; weight targets agreed and set, 180-item food frequency questionnaire at baseline and 12 months | a. 4<br>b. 158<br>c. 160<br>d. 2 | No details given |
|  | aspirin, lipid-lowering diet, personal traits unsuitable for participation in the trial |  | **Physical activity:** initial 8 weeks the intensity and duration of supervised endurance workouts increased progressively then maintained at 3 times per week for 1 hour each session at 60-80% max HR as assessed at baseline using treadmill; 60% of each workout was aerobic, 25% circuit training and 15% fast walking/jogging, attendance measured and exercise logbook kept<br>**Smoking:** all participants advised to stop smoking<br>**Control:** told not to change lifestyle and that after one year they would be offered dietary advice and supervised physical training<br>a. Diet<br>b. Exercise<br>c. Diet + exercise<br>d. Control |  |  |
| Pritchard 1999<br>Pritchard et al., 1999<br>**Location:** University general practice, Lockridge, Western Australia | **Inclusion criteria:** Either sex, 25-65 years, patients with known history of type 2 diabetes, hypertension (BP >140/90mmHg at screening plus 2 similar recordings in past medical notes) and/or overweight (BMI >25 kg/m<sup>2</sup>)<br>**Exclusion criteria:** mental illness, intellectual handicap, terminal illness, acute illness, pregnancy, taking part in other health education programmes | No details given | **Diet:** Reduce total energy intake and to reduce intake from fat to no more than 30%, CHO no less than 50% and 20% protein<br>**Counselling:** Counselling on principles of good nutrition and exercise and addressed problem areas in lifestyle and dietary patterns; counselled on food, shopping and cooking, food selection, meal planning and exercise programmes, advised to complete food records and diet history, participants discouraged from smoking and to have 2 or more alcohol free days per week with no more than 2 alcoholic standard drinks per day for females and 4 for males<br>**Control:** Participants received their usual care by GP but did not receive any counselling by dietician, mailed to reattend at 12 months | a. 7<br>b. 7<br>c. 2 | a. Dietician + Doctor<br>b. Dietician<br>c. Control |
|  |  |  | a. Dietician – diet + counselling advice<br>b. Dietician + GP (saw GP at baseline and on 2 other occasions during the 12 months)<br>c. Control |  |  |
| Swinburn 2001<br>Swinburn et al., 2001<br>**Location:**<br>University of Auckland, New Zealand | **Inclusion criteria:** Either sex, 40 years or more, IGT (OGTT, 2 hour plasma glucose 7.8-11.0 mmol/L) or high normal plasma glucose (OGTT, 2 hour plasma glucose 7.0-11.0 mmol/L)<br>**Exclusion criteria:** none stated | No details given | **Diet:** Reduced fat ad libitum diet, education and identification of strategies to reduce fat intake, personal goal setting, self-monitoring through food diaries, food label reading<br>a. Low-fat diet<br>b. Control – usual diet, general dietary advice regarding healthy food choices given at baseline only | 13 | No details given |
| Wood 1991<br>Wood et al., 1991<br>**Location:**<br>Stanford University, California, USA | **Inclusion criteria:** Either sex, 25-49 years, 120%-150% IBW, BMI 28-34 kg/m<sup>2</sup> males, 24-30 kg/m<sup>2</sup> females, non-smokers, sedentary (exercise less than twice per week, less than 30 minutes each time), resting BP <160/95 mm Hg, plasma cholesterol <6.72 mmol/L, plasma triglycerides <5.65 mmol/L, less than average 4 alcoholic drinks per day, generally good health<br>**Exclusion criteria:** medication known to affect BP or lipid metabolism, pregnancy, lactating or taking oral contraceptive in last 6 months or planning pregnancy in subsequent 2 years | No details given | **Diet:** NCEP step 1 diet consisting of 55% CHO, 30% fat with saturated fat no more than 10%, dietary cholesterol <300mg/day, calorie reduction<br>**Physical activity:** Aerobic exercise (brisk walking or jogging) at 60-80% maximum heart rate initially for 25 minutes 3 times per week increasing to 45 minutes 3 times per week by month 4, monthly activity logs kept<br>**Control:** Instructed to maintain usual diet and exercise patterns<br>a. Diet<br>b. Diet + exercise<br>c. Control | a. 25<br>b. 181<br>c. 2 | Registered dietician |

**Table S3.**
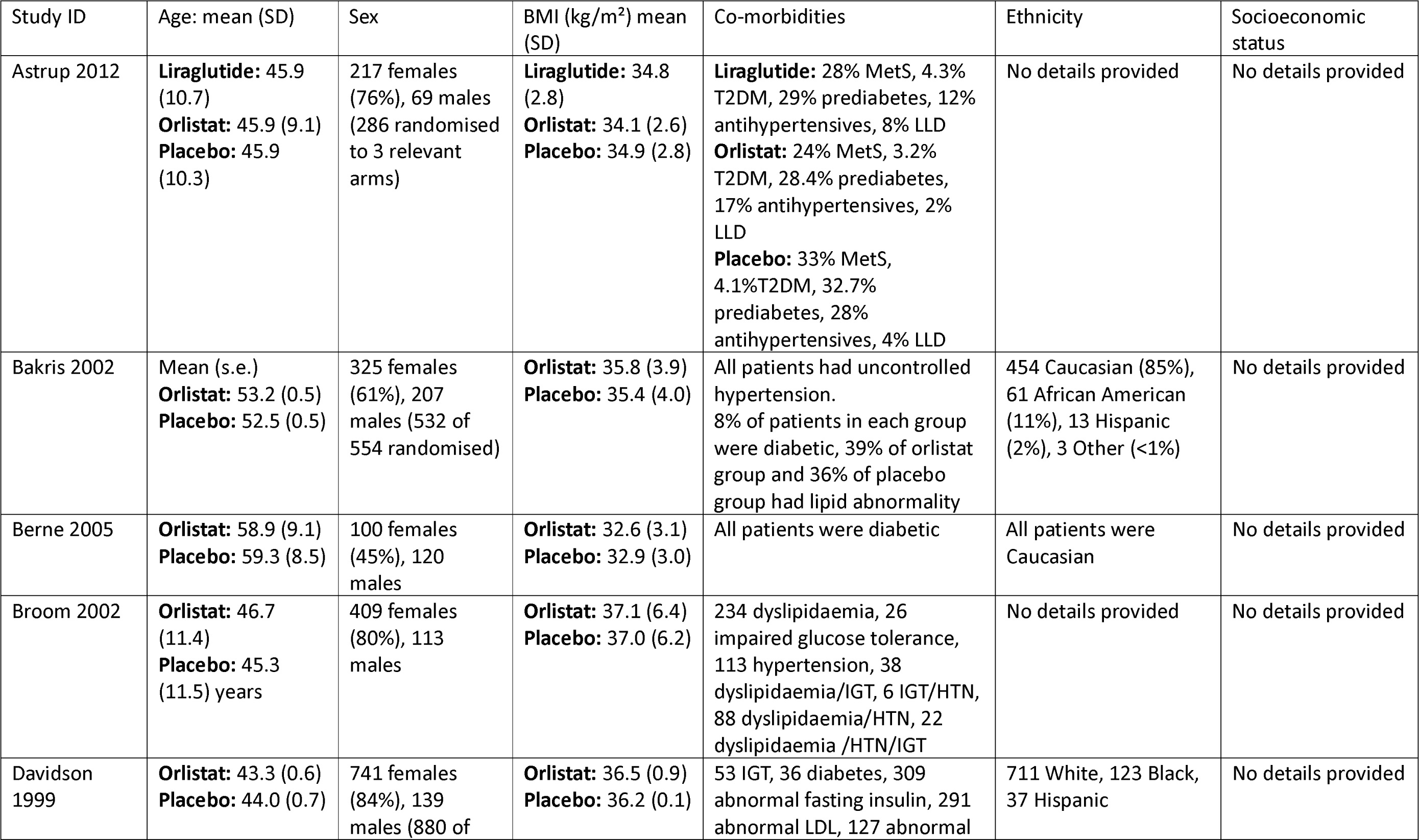
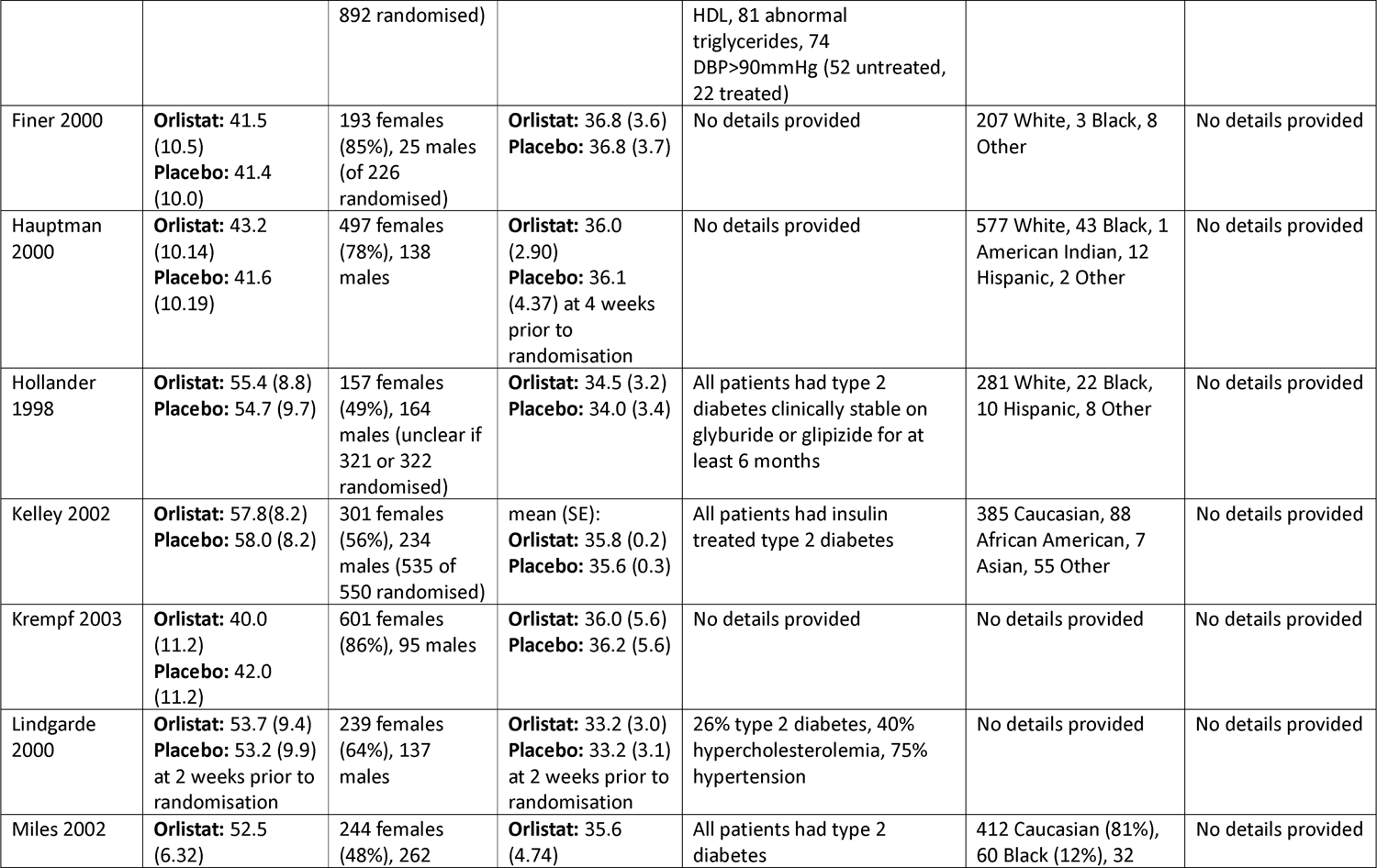
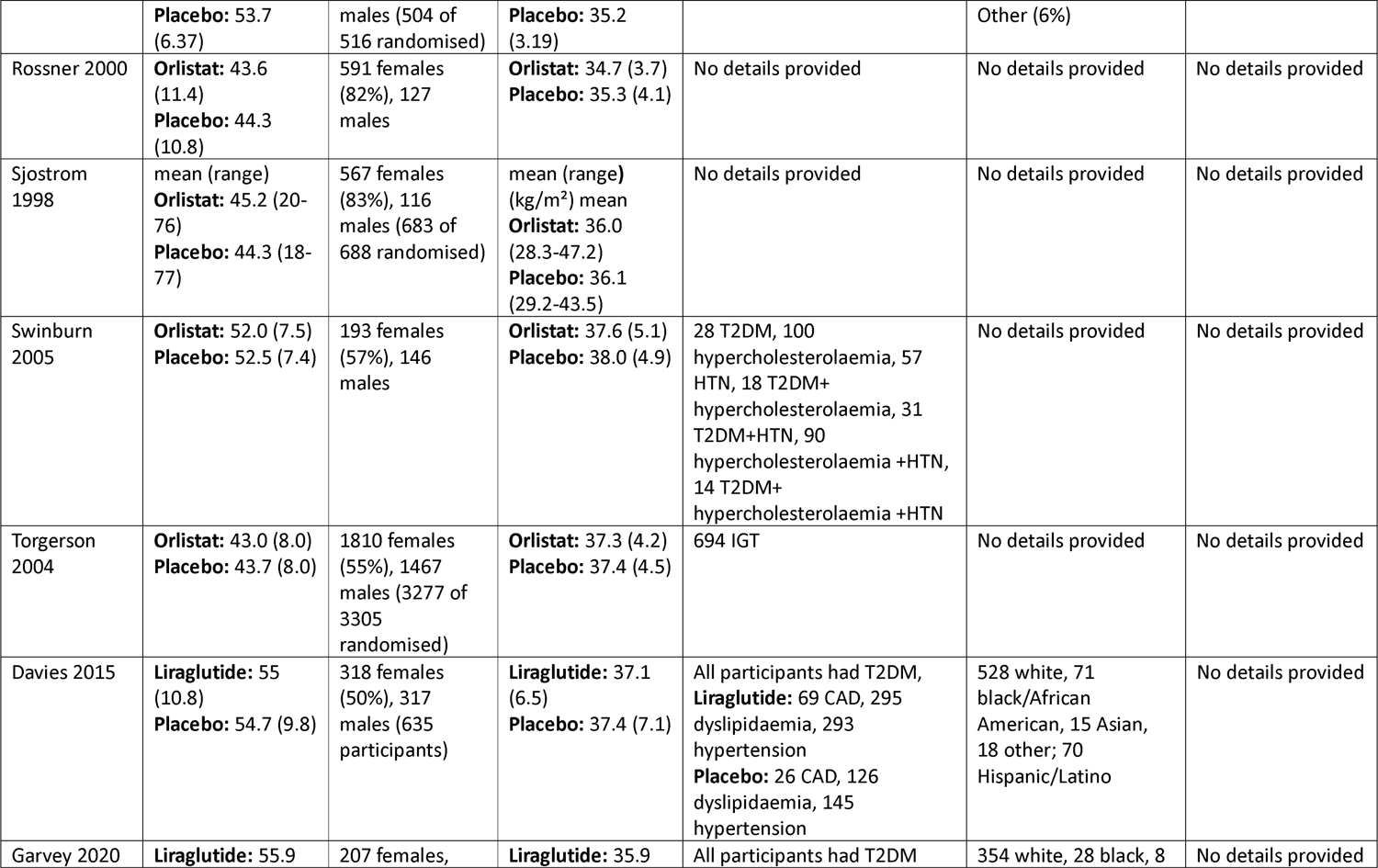
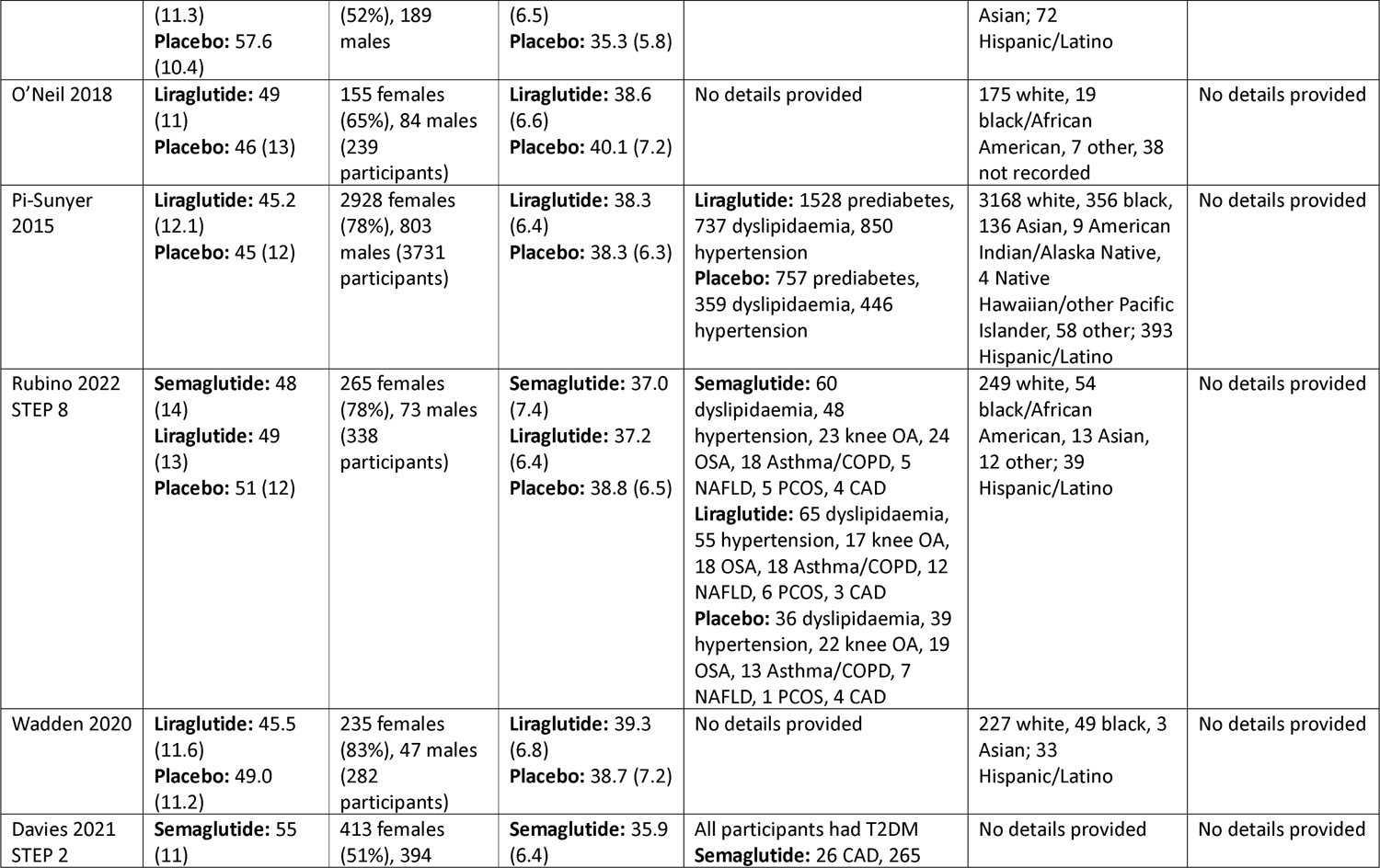
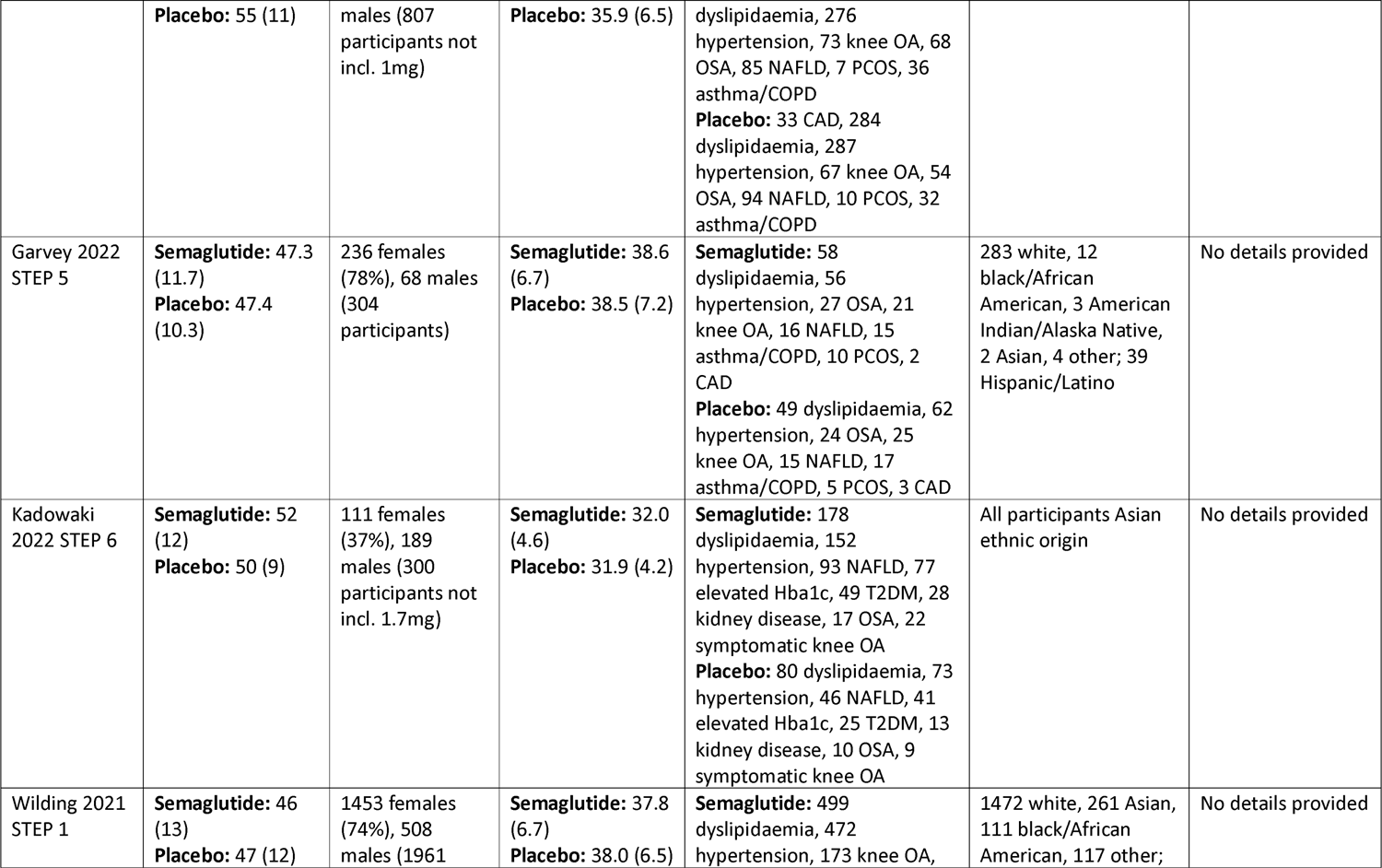
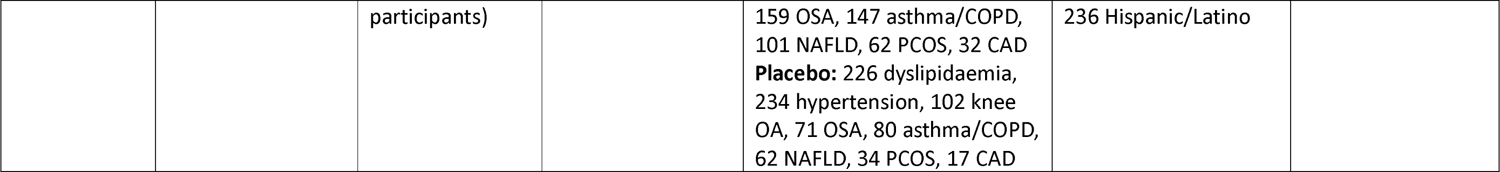
Description of drug trials’ baseline data.

**Table S4.** Description of low-fat reducing diet trials’ baseline data.

| Study ID | Sex | Age: mean (s.d.) | Co-morbidities | Ethnicity | Socioeconomic status | BMI: (kg/m<sup>2</sup>) mean (s.d.) | Citation |
| --- | --- | --- | --- | --- | --- | --- | --- |
| Abbenhardt 2013 | 439 female, 0 male | 58.0 (5.0) years<br>**Diet:** 58.1 (6.0)<br>**Control:** 57.4 (4.4) | No details provided | 372 Non-Hispanic White, 35 African American, 8 Asian/Pacific Islander, 12 Hispanic/Latino, 11 Other | No details provided<br>**Education:** 286 college degree<br>**Marital status:** 278 married or have partner<br>**Smoking:** 181 ever smoked, all current non-smokers | **Diet:** 31.1 (3.9)<br>**Control:** 30.7 (3.9) | Abbenhardt et al., 2013 |
| HOT 1999 | 53 female, 49 male | **Diet:** 57 (6)<br>**Control:** 59 (7) completers only | DBP>100mmHg | 41 African American, 61 White | No details provided | **Diet:** 34 (6)<br>**Control:** 34 (6) completers only | Jones et al., 1999 |
| Lim 2010 | 93 female, 20 male | **Diet:** 48.6 (11.3)<br>**Control:** 43.1 (10.7) | At least one CVD risk factor (blood pressure, serum lipids) | No details provided | No details provided | **Diet:** 30.5 (9.5)<br>**Control:** 33.3 (4.6) | Lim et al., 2010 |
| ODES 1995 | 21 female, 198 male | 44.9 (2.5) | No details provided | No details provided | No details provided | **Diet:** 29.54 (3.89) **Control:** 28.30 (3.15) | Anderssen et al., 1995 |
| Pritchard 1999 | 198 female, 75 male | 199 of 273 participants <50 years | 31% hypertension, 2% diabetes, 4% diabetes and hypertension | No details provided | 58% most disadvantaged quartile, 20% more disadvantaged, 20% less disadvantaged, 2% least disadvantaged<br>**Occupation:** 56% | Not stated<br>**Weight kg:** mean: **Dietician + Doctor:** 91.7<br>**Dietician:** 85.5<br>**Control:** 89.1 | Pritchard et al., 1999 |
|  |  |  |  |  | home duties, 20%<br>trade/driver/labourer,<br>6% unemployed, 14%<br>clerical/sales, 4%<br>manager/professional<br>**Relationship status:**<br>22% without partners,<br>78% married or de<br>facto |  |  |
| Swinburn<br>2001 | 35 female,<br>101 male<br>(136 of 176<br>randomised) | **Diet:** 52.5<br>(6.5) **Control:**<br>52.0 (6.7) | 44 diabetes, 19 IGT,<br>26 impaired fasting<br>glucose, 47 high<br>normal plasma<br>glucose | 97 European, 12 Maori,<br>22 Pacific Islander, 5<br>Other | No details provided | **Diet:** 29.08<br>(4.47) **Control:**<br>29.17 (4.02) for<br>completers | Swinburn et<br>al., 2001 |
| Wood 1991 | 132 females<br>132 males | 39.1 (6.4)<br>females 40.3<br>(6.3) males | No details provided | No details provided | No details provided | 27.9 (2.2)<br>females, 30.7<br>(2.2) males | Wood et<br>al., 1991 |

